# Nonuniform UV-C dose across N95 facepieces can cause 2.9-log variation in SARS-CoV-2 inactivation

**DOI:** 10.1101/2021.03.05.21253022

**Authors:** Alisha Geldert, Alison Su, Allison W. Roberts, Guillaume Golovkine, Samantha M. Grist, Sarah A. Stanley, Amy E. Herr

**Author notes:** These authors contributed equally (Alisha Geldert, Alison Su).

## Abstract

During public health crises like the COVID-19 pandemic, ultraviolet-C (UV-C) decontamination of N95 respirators for emergency reuse has been implemented to mitigate shortages. However, decontamination efficacy across N95s is poorly understood, due to the dependence on received UV-C dose, which varies across the complex three-dimensional N95 shape. Robust quantification of UV-C dose across N95 facepieces presents challenges, as few UV-C measurement tools have sufficient 1) small, flexible form factor, and 2) angular response. To address this gap, we combine optical modeling and quantitative photochromic indicator (PCI) dosimetry with viral inactivation assays to generate high-resolution maps of “on-N95” UV-C dose and concomitant SARS-CoV-2 viral inactivation across N95 facepieces within a commercial decontamination chamber. Using modeling to rapidly identify on-N95 locations of interest, *in-situ* measurements report a 17.4 ± 5.0-fold dose difference across N95 facepieces in the chamber, yielding 2.9 ± 0.2-log variation in SARS-CoV-2 inactivation. UV-C dose at several on-N95 locations was lower than the lowest-dose locations on the chamber floor, highlighting the importance of on-N95 dose validation. Overall, we couple optical simulation with *in-situ* PCI dosimetry to relate UV-C dose and viral inactivation at specific on-N95 locations, informing the design of safe and effective decontamination protocols.

## Introduction

The global shortages of N95 respirators during the COVID-19 pandemic has required crisis capacity strategies for decontamination and reuse of these complex, multilayered, made-for-single-use protective textiles. With established applications in water^1,2^, air^3,4^, and non-porous surface^5^ disinfection, ultraviolet-C (UV-C) germicidal (200-280 nm) irradiation was identified as a promising and accessible method for N95 decontamination^6^. Upon sufficient absorption by nucleic acids, UV-C inactivates pathogens by damaging their genetic material^5^; thus, UV-C decontamination efficacy is critically dependent on total received dose (integrated irradiance over time). The U.S. Food & Drug Administration (FDA) definition of tier 3 “bioburden reduction” requires sufficient UV-C dose to be applied across the N95 to yield ≥3-log_10_ inactivation of non-enveloped virus^7^ (“log_10_” subsequently referred to as “log”).

N95s present distinct challenges for UV-C decontamination: the applied surface dose required to decontaminate all N95 layers is orders of magnitude higher than the dose required on non-porous surfaces^8,9^, and varies between N95 models^10,11^ due to substantial differences in material composition. UV-C decontamination protocols must ensure that all N95 surfaces receive sufficient dose for pathogen inactivation, while also not exceeding the exposure threshold for material degradation, as high cumulative doses of UV-C degrade N95 material^12^. Additionally, UV-C dose is nonuniformly distributed across the complex, 3D N95 surface due to Lambert’s cosine law^13^ and self-shadowing. Thus, UV-C distribution across N95 surfaces is highly dependent on N95 morphology, as well as the decontamination system and N95 positioning. Together, these characteristics complicate determination of the UV-C dose applied to all N95 surfaces in a decontamination system, impacting both research and implementation^14,15^.

Effective implementation requires translation of robust research studies linking on-N95 surface dose to viral inactivation for a given UV-C source emission spectrum and N95 model. However, coincident on-N95 dose and viral inactivation measurements are infeasible as the measurement sensor would shadow the pathogen. Furthermore, most UV-C dosimetry tools lack sufficient spatial resolution, throughput, and angular response for on-N95 measurements. As a result, UV-C dose for N95 decontamination is typically characterized indirectly. For example, to circumvent challenges associated with making UV-C dose measurements on non-planar surfaces, many studies, including a recent study of SARS-CoV-2^16^, assess UV-C dose and viral inactivation on flat coupons of N95 material. N95 coupon studies determine the UV-C dose required for viral inactivation throughout the porous N95 material layers, but fail to capture the impact of the 3D facepiece shape on the received UV-C dose across the N95 surface. Other approaches use optical modeling to estimate the UV-C distribution across N95 surfaces from the UV-C dose measured in a single location, in order to relate approximate UV-C dose to SARS-CoV-2 inactivation^17^. Optical modeling is an attractive approach to study UV-C distribution, as it can recapitulate nearly any UV-C system to provide a high-resolution map of irradiance distribution^18^ via entirely user-defined system parameters. However, optical models alone cannot capture non-idealities such as irradiance fluctuations, bulb-to-bulb differences in power output, and environmental and material changes over time^5,14,19,20^. Additionally, while the modularity of optical modeling is advantageous for broad applicability, the model accuracy depends on both the optics expertise of the user and the accuracy of user-defined parameters such as the reflective and scattering properties of all materials. Thus, the high resolution and rapid iteration capabilities of optical simulations would be most valuable when coupled with *in situ* validation measurements.

To this end, a promising *in situ* method has recently been developed to quantify on-N95 dose using UV-C photochromic indicators (PCIs)^14^. PCIs complement simulation results by providing absolute dose measurements and empirical validation. Planar, paper-like dosimeters similar to PCIs have been shown to have ideal angular detection response^21^, though the angular response of UV-C PCIs has not been quantified. The low cost and small, flexible form factor of PCIs supports quantitative, spatially resolved and high-throughput on-N95 PCI dosimetry in the same exposure and in nearly the same on-N95 location as inoculated pathogens, minimizing confounding factors such as temporal or spatial variation^14^ and angular dependence of UV-C irradiance^13^. Thus, PCIs may comprise a cornerstone to better inform safe and effective UV-C decontamination, especially when corroborated by further study to confirm PCI angular response and suitability for readout by diverse, lower-cost color readers.

Here, to investigate the impact of UV-C dose variation on SARS-CoV-2 inactivation on N95s, we introduce a method to simultaneously map UV-C dose and SARS-CoV-2 viral inactivation across N95 respirator facepieces. We integrate two approaches for high-spatial-resolution on-N95 dosimetry: PCI quantification and optical modeling. We develop an optical modeling workflow to characterize UV-C dose distribution across N95s within a decontamination chamber to rapidly iterate on experimental design, and simultaneously inform and validate this model using *in-situ* PCI dose quantification. From the high-resolution simulated N95 dose maps, we identify pairs of proximal measurement sites receiving equivalent UV-C dose in order to measure UV-C dose at SARS-CoV-2 inoculation sites within the same UV-C exposure. For the first time, we apply quantitative *in-situ* PCI dosimetry to simultaneously quantify UV-C dose and SARS-CoV-2 inactivation across a model N95 facepiece (intra-N95) at multiple locations (intra-chamber), providing new, practical insight into how N95 facepiece shape impacts decontamination efficacy.

## Materials and Methods

### Inter-UV-C chamber and radiometer assessment

All UV-C decontamination experiments were performed with Spectronics XL-1000 UV-C chambers with BLE-8T254 low pressure amalgam bulbs. Irradiance was measured using calibrated, NIST-traceable ILT 1254/TD UV-C radiometers (International Light Technologies, ILT) and corresponding ILT DataLight III meter software. A custom notch in the UV-C chamber doors allowed a cable to pass through for *in-situ* radiometer measurements. One chamber and radiometer were used exclusively in a biosafety level 3 (BSL-3) laboratory for SARS-CoV-2 inactivation experiments, while another set was used for all experiments outside BSL-3. The UV-C irradiance over time and space within the two chambers were concordant (Figure S1), as were measurements from the two radiometers (Figure S2).

### PCI measurements

For UV-C dose measurements, PCIs (UVC 100 Dosimeter dots, American Ultraviolet; 25.4 mm diameter) were cut into quarters prior to use. D65/10° L*a*b* PCI color was measured using an RM200QC spectrocolorimeter (X-Rite, large aperture setting) and/or Color Muse colorimeter (Variable, Inc, with Variable color app). The Color Muse was aligned over the PCI using a template (Figure S3). PCI quantification was performed as described previously^14^, summarized in Note S1. The PCI dynamic range was defined as the dose range over which relative uncertainty was <10%. To measure doses beyond the PCI dynamic range on planar surfaces, a 1.1 mm-thick Borofloat glass attenuator (25.4 mm width and length, 80/50 scratch/dig quality, Precision Glass & Optics 0025-0025-0011-GE-CA) with 12.4% ± 0.4% UV-C transmittance (measured in a UV-C chamber) was placed over the PCI (Figure S4). We generated calibration curves specific to the PCI batch, attenuator, and colorimeter to quantify UV-C dose from PCI color change (CIEDE2000 ΔE) with respect to an unexposed reference (Figure S4, Figure S5). PCI angular response was characterized by measuring the RM200QC-measured dose quantified from PCIs exposed to a UV-C point source at different angles of incidence (Figure S6).

For all ratios of two PCI measurements (e.g., fold difference in on-N95 dose), other than cases where PCI measurements are normalized to a maximum PCI reading, we report total error: the root sum square of standard deviations associated with both replicate variation and propagated uncertainty in PCI dose estimation. All other error values report the standard deviation of replicate measurements.

### Optical model

To create a model of the respirator compatible with the optical modeling software, a 3M 1860 N95 with straps removed was scanned using a Creaform Go!SCAN 3D. After additional pre-processing, the N95 was positioned within a CAD model of the UV-C chamber (Figure S7). The entire assembly was then imported into non-sequential mode in Zemax OpticStudio (Version: 20.3) and exploded into individual parts. Parts not essential to the optical model (e.g., screws, hinges, etc.) were ignored during simulations. UV-C source and surface parameters are listed in Table S1. The N95 CAD object was converted to an absorbing detector, consistent with a previous study that approximated on-N95 UV-C distribution using an absorbing spherical detector^18^, and positioned and/or duplicated to match *in-situ* chamber locations. All simulations were performed with “Use Polarization”, “Scatter NSC Rays”, “Split NSC Rays” and “Ignore Errors” engaged. Detector data were exported and analyzed using custom MATLAB scripts. Because the optical model may not accurately predict absolute dose due to environmental fluctuations, simulation results were normalized to the maximum value within the analyzed domain (e.g., entire chamber and/or N95(s)).

### UV-C dose distribution on chamber floor

UV-C dose distribution across the chamber floor was characterized *in situ* at 15 evenly spaced locations (Figure S8) using PCIs as described previously^14^; briefly, all 15 PCIs were simultaneously exposed to ∼100 mJ/cm^2^, then read with the RM200QC within 600 s. Peak UV-C irradiance within a 15 s exposure was also measured at 14 of these locations sequentially using a radiometer (the built-in chamber sensor obstructed placement at one location). Simulated UV-C dose at each location was extracted from the optical model using custom MATLAB scripts.

### UV-C dose distribution across N95 facepieces

#### In situ

To empirically measure on-N95 UV-C dose, PCIs with backing removed were adhered to the N95 facepiece, exposed, and subsequently removed for color quantification. To facilitate comparison to simulation, each PCI location on the N95 was recorded by measuring the PCI: 1) corner height (C), 2) highest point height if not corner height (lowest point height if corner is highest) (h), 3) rotation along the N95 surface (Φ, Figure S9), and 4) lateral distance from either the nosepiece-to-chin midline or side-to-side seam. N95 straps were removed to minimize shadowing and variability in N95 tilt. A printed floor map ensured reproducible N95 positioning in the chamber, with nosepieces toward the door (Figure S10).

#### Optical model

To characterize on-N95 UV-C dose from simulations, average values at specific *in-situ* PCI locations were extracted from the N95 detector simulation data using a custom MATLAB script. Briefly, N95 detector data were imported into MATLAB. The outline of each PCI was plotted on top of the simulated N95 dose map using the spatial parameters described above. The vertical height of the PCI (d) was then defined as |C-h|. The angle of rotation toward the N95 surface (α, Figure S9) of the PCI was calculated based on geometry. When the corner is either the highest or lowest part of the PCI:

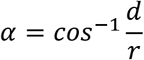

where *r* is the radius of the PCI.

When corner is not the highest or lowest point of the PCI,

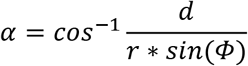

where Φ is defined as the angle between the horizontal axis and highest PCI point.

The average on-N95 value from simulation was calculated as the mean value of the data points contained within the PCI perimeter, determined using the “inpolygon” function on a 2D projection of N95 data points and PCI outlines (Figure S9).

### Heatmap plots

All heatmaps of UV-C dose or irradiance were generated with the ‘inferno’ perceptually uniform, colorblind-friendly colormap, which was created by Stéfan van der Walt and Nathaniel Smith and adapted from Python’s matplotlib for use in MATLAB by Ander Biguri.^22^

### SARS-CoV-2 preparation and handling

For all viral inactivation experiments, SARS-CoV-2 preparation, stock titration, inoculation, and TCID_50_ assay were performed similarly to previous studies^23^ (Note S2). In brief, 3 aliquots of 16.67 μL (50 μL in total) of SARS-CoV-2 stock at 8 × 10^7^ TCID_50_/mL were loaded per N95 inoculation site and dried for 3.5 hours at room temperature in a biosafety cabinet. After UV-C exposure, inoculation sites were excised with a 12 mm-diameter biopsy punch (MedexSupply ACD-P1250), incubated in DMEM supplemented with 10% fetal bovine serum, 100 U/mL penicillin, and 100 μg/mL streptomycin for ≥30 minutes, and viable SARS-CoV-2 virus was quantified by a TCID_50_ assay. All study procedures were approved by the UC Berkeley Committee for Laboratory and Environmental Biosafety and conducted in agreement with BSL-3 requirements.

### SARS-CoV-2 dose response on N95 coupons

The UV-C dose response of SARS-CoV-2 was assessed by measuring viral inactivation on 3M 1860 N95 coupons inoculated with SARS-CoV-2 and exposed to different UV-C doses. By mapping UV-C irradiance across the chamber floor, we identified 5 locations of equivalent irradiance at which to place a radiometer, PCI, and 3 inoculated N95 coupons (Figure S11, Figure S12). PCIs and coupons were placed on custom-built platforms to match the height of the radiometer sensor. Platforms were built from laser-cut (HL40-5G-110, Full Spectrum Laser) pieces of 3.175 mm-thick acrylic (McMaster Carr 85635K421), joined with epoxy (J-B Weld 50176). Printed maps on the chamber floor and platforms ensured consistent positioning from run-to-run (Figure S13).

For SARS-CoV-2 inactivation experiments, 3 replicate inoculated coupons were simultaneously irradiated with a given UV-C dose. Given the minimal impact of expiration status on UV-C decontamination efficacy (Figure S14), expired (i.e., past the manufacturer-recommended shelf life) N95s were used for experiments, to preserve non-expired N95s for healthcare workers. Coupons (15 mm × 20 mm) were cut from the edge of N95s to include the raised, sealed seam to minimize layer separation. The seam did not prevent the coupons from lying flat during UV-C exposure. Both a radiometer and PCI (with Borofloat attenuator for doses beyond the PCI upper limit of quantification) were used to quantify *in-situ* UV-C dose applied during each exposure. Exposure time was estimated by dividing the target dose by the irradiance at the coupon platform (∼6.4 mW/cm^2^ after bulb warm-up). To account for output degradation^14^ (Figure S1), exposure time was optimized by comparing the dose measured by the radiometer during a test exposure to the target dose.

After each exposure, PCI(s) were measured with both the RM200QC and the Color Muse. Biopsies were excised from all irradiated coupons, as well as one unexposed control coupon stored at room temperature outside the UV-C chamber. A TCID_50_ assay was performed to assess SARS-CoV-2 viability (Note S2).

### Paired measurements of UV-C dose and SARS-CoV-2 inactivation on N95s

To simultaneously measure UV-C dose and SARS-CoV-2 inactivation on 3M 1860 N95 facepieces, PCIs (without attenuator) were affixed to N95s at each chosen dose measurement site, and accompanying SARS-CoV-2 inoculation sites outlined in advance to facilitate accurate viral deposition. SARS-CoV-2 was inoculated at each paired inoculation site. After drying, two N95s (‘corner’ and ‘front’ N95s) were placed in a UV-C chamber after bulb warm-up. To monitor dose during each exposure, a radiometer and PCI were also placed at their respective positions near the two corners of the chamber floor (Figure S10). After a 10 s UV-C exposure, PCIs were removed from the N95s and measured with both the RM200QC and the Color Muse. SARS-CoV-2 inoculation sites as well as an unexposed room temperature control coupon were excised following each UV-C exposure.

## Results and Discussion

### Measuring UV-C dose and SARS-CoV-2 inactivation on and across N95s

In this study, we sought to understand the impact of N95 shape and placement on SARS-CoV-2 inactivation and how variation in inactivation relates to UV-C dose received across the N95 surfaces (Figure 1A). Building upon previous work quantitatively relating PCI color change to received UV-C dose^14^, we introduce simultaneous measurement of SARS-CoV-2 inactivation and UV-C dose on N95 facepieces. We extend characterization of the quantitative PCI dosimetry method^14^ in two ways. First, we measured the angular response of PCIs and verified a near-ideal response (Figure S6), confirming that PCIs are suitable to measure UV-C dose on non-planar surfaces. Second, to increase accessibility of the PCI dosimetry method, we compared the performance of a substantially lower-cost colorimeter to the previously reported spectrocolorimeter (Figure S4). Applying this PCI dosimetry method, we paired PCI UV-C dose measurements with SARS-CoV-2 inactivation measurements to characterize the received dose and resulting viral inactivation variation across N95 facepiece surfaces.

**Figure 1.**
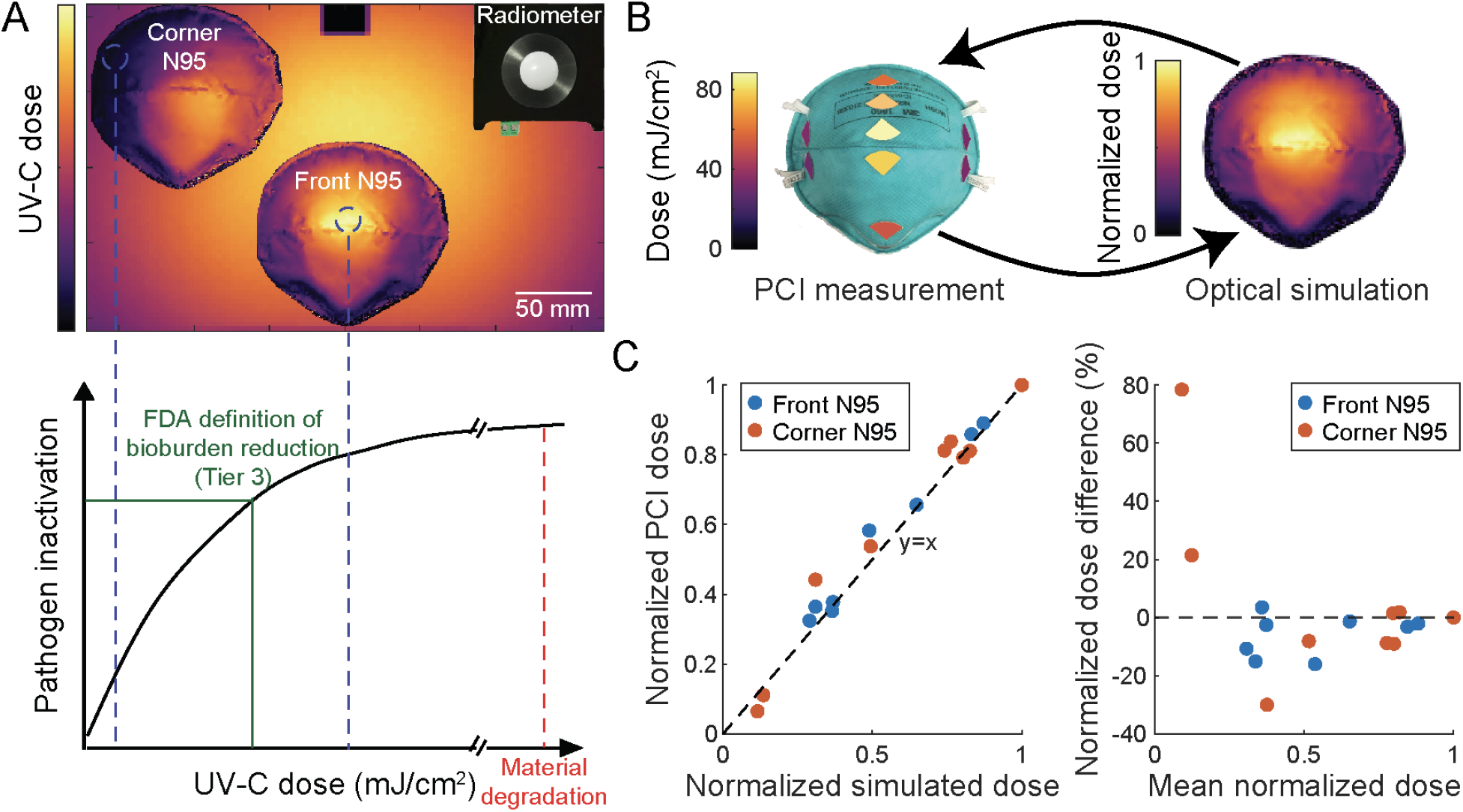
Integrated optical modeling and *in-situ* PCI measurement pipeline for simultaneous and near-coincident on-N95 UV-C dose and viral inactivation measurements. (A) Schematic highlighting how UV-C dose received across complex N95 surfaces can vary substantially, creating a narrow range of UV-C doses that deliver sufficient dose for pathogen inactivation while not exceeding the exposure threshold for material degradation. (B) *In-situ* PCI measurements and optical simulation results were used in tandem to inform and rapidly iterate on experimental design. (C) Comparison of normalized *in-situ* PCI and simulated doses at seven discrete locations on N95s in two different chamber positions. Normalized dose difference was calculated as 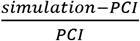. Black lines (y = x on left, y = 0 on right) indicate where the data would lie if PCI and simulation measurements were equal. Simulation tends to overestimate normalized *in-situ* PCI dose at low doses and underestimate *in-situ* PCI dose at high doses.

Towards the study goal of assessing impact of N95 shape and placement on received UV-C dose and SARS-CoV-2 inactivation, we first identified the dynamic range of the UV-C dose-response curve. Only doses within this range will elucidate the variable relationship between UV-C dose and viral inactivation. The physical setup and exposure time of N95s in a decontamination chamber and the SARS-CoV-2 inoculation sites were then optimized to receive doses spanning that dynamic range. To perform this non-trivial optimization, we implemented both *in silico* optical ray-trace modeling and *in-situ* experimental PCI quantification. We iterated between high-resolution modeling predictions and the more accurate on-N95 PCI-based UV-C dose measurements (Figure 1B).

### Building and validating optical model of a UV-C decontamination system

After optically modeling the UV-C decontamination chamber, we observed normalized simulation dose measurements differ from normalized *in-situ* radiometer measurements by an average of 4.7% ± 4.5% at 14 unique locations across the chamber floor (Figure S11). Assuming spatially invariant fluctuations, the normalized irradiance and normalized dose distribution within the system are equal. Therefore, the terms “normalized irradiance” and “normalized dose” are used interchangeably to compare to *in-situ* results, depending on the *in-situ* measurement approach (i.e., radiometer or PCI). From a 3D scan of a 3M 1860 N95 imported into the virtual UV-C chamber, normalized simulation measurements differ from normalized *in-situ* PCI measurements by an average of 6.0 ± 6.2% across the facepiece centrally positioned near the chamber door (‘front N95’) (Figure 1C). The largest discrepancy on the door-facing N95 surface saw simulation underestimate the normalized PCI dose by ∼16%. For an N95 positioned in the chamber rear corner (‘corner N95’), simulation differed from *in-situ* PCI measurements by an average of 18 ± 25%, with the largest discrepancies again occurring on the wall-facing N95 surfaces (Figure 1C and Figure S15). Differences between the simulation and *in-situ* measurements may arise due to N95-to-N95 shape variability (Figure S16), differences between true and modeled surface properties, and higher relative uncertainty of low-dose PCI measurements. Overall, however, on-N95 dose measurements correlate with simulation measurements at corresponding locations (Figure 1C). Thus, after validating the agreement between the simulation and *in-situ* measurements across both the chamber floor and an N95 in multiple chamber locations, we coupled the two measurement tools to design and optimize paired UV-C dose and SARS-CoV-2 inactivation experiments, leading to the first demonstration of simultaneous on-N95 viral inactivation and UV-C dose measurements to date.

### Establishing dose-response for SARS-CoV-2 viral inactivation by UV-C

In order to quantify the UV-C dose dependence of SARS-CoV-2 viral inactivation without the added complexity of the N95 facepiece shape, we first considered SARS-CoV-2 viral inactivation using UV-C on coupons of N95 material. Simulation and *in-situ* measurements identified and validated five locations in a UV-C decontamination chamber that receive equivalent UV-C irradiance (<5% variation, Figure 2A) for location-paired UV-C dose and SARS-CoV-2 inactivation measurements on N95 coupons. We simultaneously exposed triplicate coupons inoculated with SARS-CoV-2 while recording the applied dose using both a radiometer and a PCI (Figure S17). As the dynamic range of the PCIs measured with either color reader was insufficient to measure UV-C doses >∼260 mJ/cm^2^ (Figure 2B), for these higher doses we placed 1.1-mm thick Borofloat glass over the PCI on the flat PCI platform to attenuate incident UV-C irradiance and extend the PCI dynamic detection range. We observed an extended upper limit of quantification (ULOQ) of 1853.2 mJ/cm^2^ for the Borofloat-PCI pair when PCI color is measured with the spectrocolorimeter, compared to 261.4 mJ/cm^2^ without the attenuator (Figure S4). When using the lower-cost colorimeter, Borofloat extended the ULOQ from 168.1 mJ/cm^2^ to 802.6 mJ/cm^2^ (Figure S4). While the ULOQ of the Borofloat-PCI pair when using the lower-cost colorimeter was lower than some of the UV-C doses included in the SARS-CoV-2 dose-response measurements, we observed good agreement in estimated dose using both color readers to measure all PCIs in SARS-CoV-2 experiments (Figure S4). Borofloat was only paired with PCIs on planar surfaces (not on-N95), as Borofloat transmittance may depend on incident angle, yielding non-ideal angular response.

**Figure 2.**
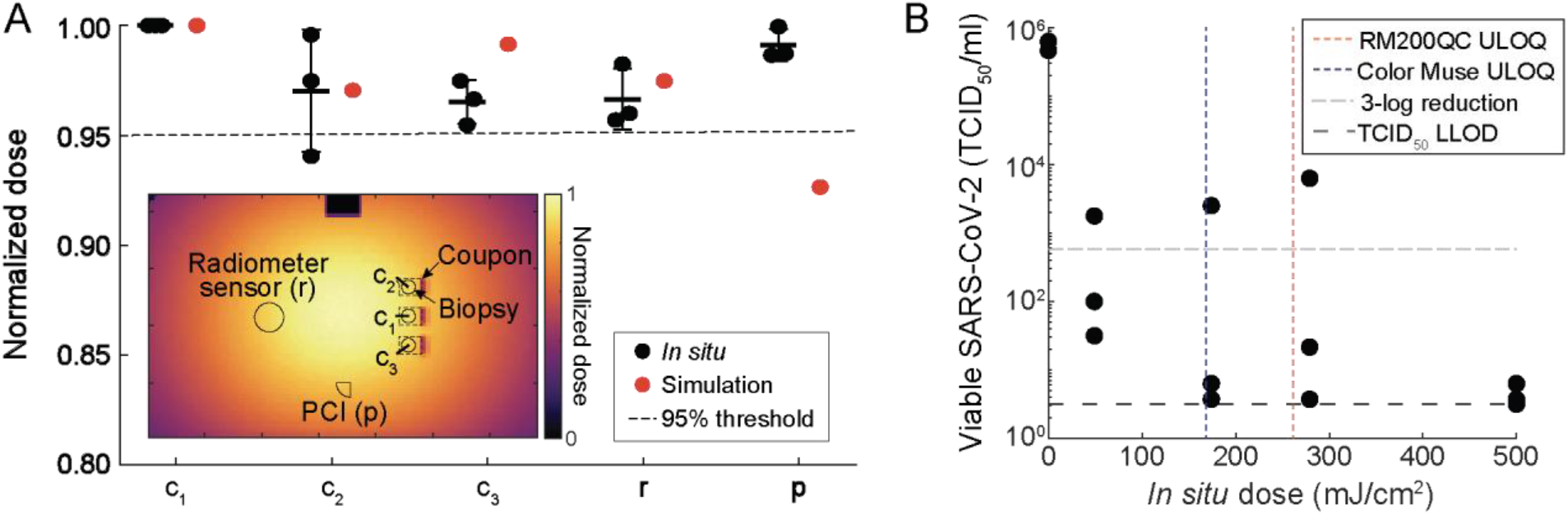
Measurement of UV-C dose required for SARS-CoV-2 inactivation on N95 coupons is informed by optical modeling and *in-situ* PCI dose measurements. (A) Using the optical model and validating *in situ*, five locations within the decontamination chamber receiving similar UV-C doses (<5% variation between mean *in-situ* dose measurements at each location) were identified. To inform biopsy location, the optical model also assessed the impact of each coupon seam (modeled as 15 mm wide × 2.5 mm tall × 1 mm thick absorbing rectangular volumes at the right-hand side of each coupon) on UV-C distribution. *In-situ* measurements were made using PCIs to simultaneously measure dose received at the PCI and 3 coupon locations while simultaneously recording irradiance with the radiometer. Mean and standard deviation are indicated for the *in-situ* measurements. (B) SARS-CoV-2 recovery on N95 coupons is dependent on *in-situ* UV-C dose, measured using a radiometer. During UV-C exposure, the radiometer, PCI, and triplicate N95 coupons were each placed as shown in (A). A Borofloat attenuator was placed on top of PCIs to measure doses >168 mJ/cm^2^ due to the limited PCI ULOQ. N=3 replicates per dose. ULOQ = upper limit of quantification. LLOD = lower limit of detection.

To elucidate the SARS-CoV-2 dose-response curve, we measured SARS-CoV-2 viral activity from N95 coupons after exposure to applied UV-C doses ranging from 500-1500 mJ/cm^2^. The applied UV-C range was selected based on previous results demonstrating that ≥1000 mJ/cm^2^ UV-C dose is required for 3-log inactivation of SARS-CoV-2 analogs on the majority of N95 models tested^10,24,11^. For all replicates exposed to 500-1500 mJ/cm^2^, we observed >5-log SARS-CoV-2 reduction on N95 coupons (Figure S18). Furthermore, any remaining virus was below the limit of detection of the TCID_50_ assay for all but one replicate, signifying that lower doses are required to identify the dynamic range of the dose-response curve of our assay.

We next assessed an applied UV-C dose range of 50-500 mJ/cm^2^. For these lower UV-C doses, we observed an average of >3-log reduction of viable SARS-CoV-2 virus at all doses (Figure 2B), with no significant differences observed between non-zero UV-C dose conditions (N = 3 replicates, Kruskal-Wallis test with Dunn’s multiple comparison test). We also confirmed that any temperature increase in the chamber during exposure does not contribute to SARS-CoV-2 inactivation (Note S3, Figure S19). We postulate that the >20x higher SARS-CoV-2 UV-C susceptibility observed in this study as compared to previous literature is likely attributable to two factors. First, SARS-CoV-2 was inoculated without a soiling agent (e.g., sweat or sebum surrogates); soiling agents can decrease UV-C inactivation by 1-2 logs.^25,11^ Second, the 3M 1860 N95 material was very hydrophobic (water contact angle >90°, Figure S20), and deposited viral samples ‘beaded’ on the facepiece surface. Greater UV-C decontamination efficacy has generally been observed on hydrophobic N95 models^11^, which we hypothesize may be due to the greater proportion of virus inoculated on the outer N95 layers. Because the outer N95 layers receive more UV-C dose than inner layers^9^, inactivation on hydrophobic N95s may more closely resemble nonporous surface decontamination, on which lower UV-C doses (∼4.3 mJ/cm^2^) have been shown to yield >3-log reduction of SARS-CoV-2^26^. Droplet imbibition into porous matrices is a complex process that depends on properties of the fluid and substrate^27^, differences in inoculation volume and solution, and N95 material, all of which may influence the proportion of virus which penetrates into inner N95 layers. Thus, the system scrutinized here is an idealized model system and the SARS-CoV-2 dose response behavior observed is not anticipated to represent SARS-CoV-2 inactivation in clinical settings, where different N95 models and soiling are expected to substantially increase the UV-C dose necessary. Given the results and precision of the TCID_50_ assay (Figure 2B), and the anticipated single-or two-stage exponential inactivation of virus with increasing dose^5,16,28^, we expect the dynamic range of our measured dose-response curve to exist between 0-50 mJ/cm^2^. Thus, we aimed to deliver UV-C doses within this range to map SARS-CoV-2 inactivation differences and UV-C dose nonuniformity to the complex 3D geometry of N95 facepieces (i.e., comparing among facepiece locations).

### On-N95 UV-C mapping informs design of near-coincident UV-C dose and SARS-CoV-2 inactivation measurements

Having established the UV-C dose response of SARS-CoV-2 on flat N95 coupons, we next investigated the magnitude of N95 shape-induced UV-C dose variation, as received UV-C is dependent on incident angle and distance from the UV-C source. Concomitantly, we sought to understand how the nonuniform on-N95 UV-C dose translated to SARS-CoV-2 viral inactivation efficacy. We aimed to map SARS-CoV-2 inactivation differences and UV-C dose nonuniformity across the N95 facepiece by simultaneously quantifying on-N95 dose with *in-situ* PCI measurements and SARS-CoV-2 inactivation via TCID_50_. The tandem approach allowed simultaneous measurement at multiple locations on intact N95 facepieces: a measurement not feasible with radiometers or viral inactivation measurements alone.

Because a PCI placed at the SARS-CoV-2 inoculation site would shadow the virus inoculum, we used optical simulation to identify pairs of adjacent measurement sites on-N95 which receive equal dose. With paired measurement sites, the UV-C dose received by a SARS-CoV-2 inoculation site can be monitored using a PCI placed at the proximal equivalent-dose site. Optical simulation rapidly reports the irradiance distribution across easily tunable N95 configurations with high spatial resolution, facilitating identification of: (1) *locations* to make on-N95 measurements that sample the range of delivered UV-C doses, and (2) *measurement sites* within each location receiving the same dose. Each location must be large enough to house two proximal measurement sites each ∼13 mm in diameter.

We first used optical simulation to characterize the UV-C dose distribution across the surface of multiple N95s within the chamber. To increase decontamination system throughput, multiple N95s are often irradiated simultaneously^29,30^, but care must be taken to ensure all N95s receive sufficient dose. Additionally, N95s must be separated to prevent cross-contamination. In the studied decontamination system, three N95s can be staggered within the chamber (e.g., two in the back, one in the front). Given the lateral symmetry in dose distribution within the chamber (Figure S1, Figure S11), we characterized UV-C dose distribution across two N95s in the unique positions in this ‘maximal-throughput’ layout, which we call ‘front’ and ‘corner’ (Figure 3A). From the simulated UV-C dose map across these N95s, we identified six discrete locations (a-f in Figure 3A) which sample the dose range. At locations a, b, d, and e, UV-C dose measured with PCIs *in situ* is 3.3% ± 7.6% greater than simulated dose (Figure 3A). At location f, *in-situ* UV-C dose is 46.4% ± 7.6% lower than simulated dose, in line with our previous findings that simulation overestimated *in-situ* dose by 78% near that location (Figure S15). Similarly, the largest difference between simulated and *in-situ* UV-C dose is at location c, where simulated dose is 26% ± 3% lower than the *in-situ* dose, consistent with our previous finding that simulation underestimated the *in-situ* dose by 16% near the nosepiece of the front N95 (Figure S15). The discrepancy between simulation and *in-situ* UV-C dose measurements at select on-N95 locations highlights the importance of complementary *in-situ* measurements.

**Figure 3:**
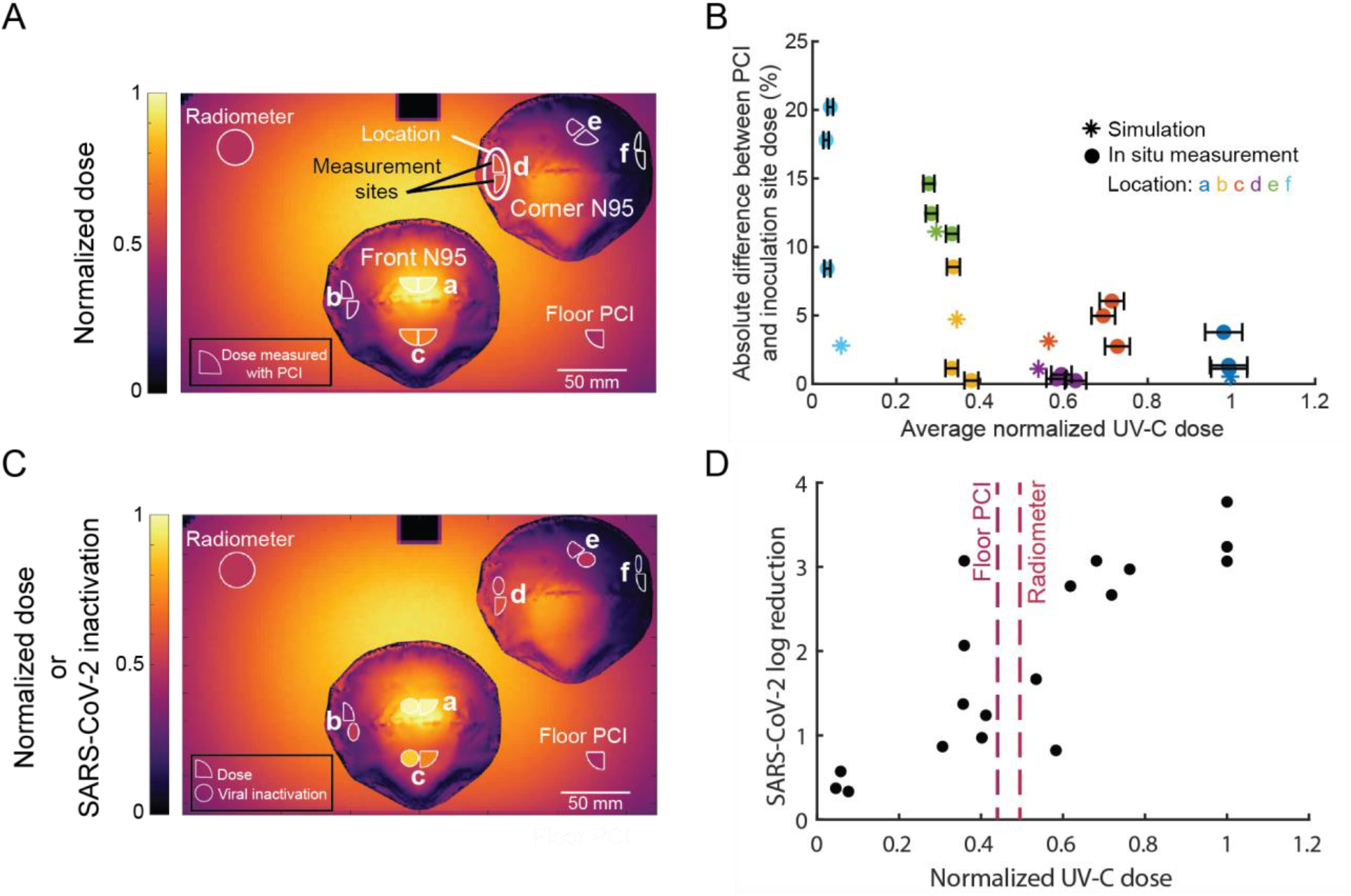
Paired on-N95 measurements of UV-C dose and SARS-CoV-2 inactivation show correlated, several-fold variation in dose and inactivation across one decontamination chamber. (A) Optical simulation of UV-C dose distribution over two 3M 1860 N95 facepieces in the UV-C chamber, overlaid with PCIs at paired measurement sites for viral inactivation and dose measurement. Heatmap shows simulated UV-C dose (normalized to the maximum dose in the chamber). PCI fill color represents the mean dose measured with PCIs *in situ* across triplicate measurements. (B) Comparison of dose differences within paired measurement sites. Data are colored by on-N95 location. Horizontal error bars on measured values represent the error in estimated dose. (C) Average normalized UV-C dose (quarter-circles) and SARS-CoV-2 inactivation (circles) at measured locations on front and corner N95, colored by the normalized value. Values are normalized to measurements at the apex of the front N95. Surrounding heatmap shows simulated UV-C dose. (D) SARS-CoV-2 inactivation on N95 facepieces is proportional to UV-C dose received. Selected locations on two N95 facepieces in the Spectronics XL-1000 UV-C chamber receive a 17.4 ± 5.0-fold difference in UV-C dose, which yields an 8.2 ± 1.4-fold difference in SARS-CoV-2 log reduction.

Within each location, the high-spatial-resolution map of simulated dose was used to identify two proposed measurement sites; dose at each site was then measured *in situ* using PCIs. Note that while measurement sites are often proximal to one another, the high-resolution simulation results established that some measurement sites receive the most similar dose when slightly offset (e.g., the sites at location b) due to the irregular N95 facepiece geometry and the off-center positioning of the N95s in the chamber. We observed that for locations receiving normalized UV-C doses >0.34 (normalized to the maximum on-N95 dose), the doses across each proximal pair of PCI and inoculation sites were within 6.0% of each other, both in simulation and when measured *in situ*. For normalized UV-C doses ≤0.34 (at more steeply sloped and/or shadowed locations), the simulated doses at each pair of measurement sites were within 11.1% of each other and the doses measured *in situ* were within 11.8% ± 6.0% of each other (Figure 3B). Differences between paired sites may be larger at locations with greater curvature (e.g., location e), where PCI angle (and thus, received UV-C dose) is more sensitive to run-to-run variation in PCI placement as well as N95 morphology. At locations with normalized UV-C doses ≤0.34, the higher relative uncertainty of PCI quantification at low doses may also contribute to a greater difference in dose at proximal sites. We quantified a relative uncertainty of ∼20% at the lowest-dose (∼5 mJ/cm^2^) location, compared to a relative uncertainty of ∼5% at all other locations when PCIs are measured with the RM200QC (Figure S4B).

### Intra-chamber variation in UV-C dose and SARS-CoV-2 inactivation

Having identified paired on-N95 measurement sites receiving equivalent dose, simultaneous measurements of UV-C dose and SARS-CoV-2 inactivation on intact N95s could be performed. We assessed N95s placed at the front and corner positions in the decontamination chamber and chose an exposure time such that dose received across the N95 surfaces would span the dynamic range of 0-50 mJ/cm^2^ determined from the coupon study (Figure 2B). For analysis, both UV-C dose and SARS-CoV-2 log reduction were normalized to the respective maximum value measured in the system within each replicate UV-C exposure.

UV-C dose and SARS-CoV-2 log reduction correspond well (Figure 3C) and are positively and linearly correlated (r^2^ = 0.7016; p = 1.4428 x 10^−5^) (Figure 3D). SARS-CoV-2 dose response is still being investigated, but is expected to be primarily log-linear^31,28,32,33^ in agreement with other pathogens. While the dose response curve likely has shoulder and/or tailing behavior at the lower and upper ends^5^, these nonlinear regions may not be captured with the range and resolution of UV-C doses tested here. The dose required for 90% inactivation (D_90_) estimated from a linear regression on the dose-response curve (r^2^ = 0.78) is ∼19 mJ/cm^2^, higher than the D_90_ of ∼1.4 mJ/cm^2^ for dried SARS-CoV-2 on a nonporous surface^31^, as expected (Figure S21).

Similar to the coupon study, we observe varying SARS-CoV-2 inactivation among replicate inoculation sites receiving similar UV-C dose (1.1 ± 0.8-log difference in inactivation between replicates), which we hypothesize may be due to: (1) the quantal nature of the TCID_50_ assay^34,35^, and/or (2) variability in the slope of the coupon surface caused by separation of N95 layers along the three sides without a seam (Figure S20). Slight variations in the amount of virus inoculated, viral extraction efficiency, and excision area may also contribute to technical variation in measured TCID_50_/mL. To characterize intra-chamber variation, we quantified the fold difference in UV-C dose and SARS-CoV-2 log reduction across both N95 facepieces in the chamber. Simulation predicted a 14.9-fold difference in UV-C dose across the facepieces of both N95s in the chamber, and we measured *in situ* a dose difference of 17.4 ± 5.0-fold. This UV-C dose range yielded an 8.2 ± 1.4-fold difference in SARS-CoV-2 log reduction (from 0.4 ± 0.1-log reduction at location f to 3.4 ± 0.4-log reduction at location a). The observed 2.9 ± 0.2-log difference in SARS-CoV-2 survival across N95s within one chamber is substantial, given the FDA definition of “bioburden reduction” on N95 respirators that requires ≥3-log reduction of various pathogens^7^. To our knowledge, this is the first study to rigorously quantify both UV-C dose and viral inactivation at paired locations on intact N95s, to understand how UV-C dose distribution and resulting decontamination efficacy depend on N95 facepiece shape.

Because *in-situ* dose is often monitored at an off-N95 location in decontamination protocols^36^, we also investigated whether dose monitoring at low-irradiance locations on the chamber floor could directly indicate the lowest on-N95 UV-C dose (Note S4). The doses in the corners of the chamber floor were 49.5% ± 1.6% (radiometer location) and 44.0 ± 0.7% (floor PCI location) of the maximum on-N95 dose, whereas the lowest on-N95 dose measured was 6.0% ± 1.6% of the maximum on-N95 dose (Figure 3C,D).Thus, in the UV-C chamber tested here, dose monitoring on the chamber floor cannot serve as a proxy for the lowest on-N95 UV-C dose.

### Intra-N95 variation in UV-C dose and SARS-CoV-2 inactivation

In addition to characterizing intra-chamber variation, we also analyzed UV-C dose and SARS-CoV-2 inactivation variation across each individual N95. On the front N95, the apex (location a) receives the highest dose while the more steeply sloped regions near the base of the sides of the N95 (location b) receive some of the lowest doses that can be measured with our approach, given the footprint of the PCI and SARS-CoV-2 inoculation site. Across the locations sampled on the front N95, simulation predicted a 3.0-fold difference in UV-C dose, and we measured a 2.8 ± 0.4-fold difference in UV-C dose using PCIs *in situ*. This variation in UV-C dose yielded a 2.8 ± 1.5-fold difference in SARS-CoV-2 log reduction (from 1.6 ± 1.2-log reduction at location b to 3.4 ± 0.4-log reduction at location a). While placing the N95 directly in the center of the UV-C chamber rather than offset toward the door would increase UV-C dose uniformity, throughput may be reduced, as the number of N95s that could fit in the chamber without contacting each other would be reduced from three to one.

Across the 3 measured locations on the facepiece of the corner N95, simulation predicted an 8.1-fold difference in UV-C dose and we measured a 10.2 ± 3.3-fold difference in dose. This variation in UV-C dose yielded a 4.9 ± 1.3-fold difference in SARS-CoV-2 inactivation (from 0.4 ± 0.1-log reduction at location f to 2.1 ± 0.7-log reduction at the maximum-dose location on the corner N95, which was either location d or e depending on the replicate). However, because the measurement locations were chosen to evenly sample the range of UV-C doses applied across both (front and corner) N95s, the measured locations on the corner N95 did not capture the maximum corner N95 dose near the apex (Figure 3A,C). Thus, we expect that the total variation in UV-C dose and resulting viral inactivation on the corner N95 would be even higher than measured here.

We also compared the magnitude of variation in UV-C dose and SARS-CoV-2 inactivation on the front and corner N95s. As compared to the front N95, the corner N95 had greater intra-N95 variation in both UV-C dose and SARS-CoV-2 inactivation. In contrast to the front N95, which had an equal amount of variation (2.8-fold) in UV-C dose and SARS-CoV-2 inactivation, the difference in UV-C dose (10.2-fold) was greater than the difference in SARS-CoV-2 inactivation (4.9-fold) on the corner N95. We hypothesize that the corner N95 receives UV-C doses which may be in the shoulder of the SARS-CoV-2 survival curve, where SARS-CoV-2 inactivation is not fully log-linear with dose^5^. If the corner N95 receives UV-C doses in this shoulder region, the magnitude of intra-N95 UV-C dose variation will be larger than the amount of variation in SARS-CoV-2 inactivation.

Characterization of UV-C dose distribution across N95s within a decontamination system is valuable for informing decontamination protocols and evaluating throughput. In our system, we observed substantially lower variation in UV-C dose and SARS-CoV-2 inactivation across a single N95, as compared to across both N95s in the chamber, which suggests that approaches to increase decontamination throughput should be carefully considered. Including more N95s in the chamber may not necessarily increase throughput as compared to a single N95 in the center of the chamber, as multiple N95s likely have more nonuniform on-N95 dose because they are more spread out and can shadow each other. Greater UV-C dose nonuniformity increases the exposure time needed for all N95 surfaces to reach the minimally acceptable UV-C dose, which in turn affects the total number of safe reprocessing cycles prior to N95 material degradation (Figure 1A)^12^. The simulation and *in-situ* dose measurement workflows we demonstrate here help inform N95 positioning within decontamination systems to optimize decontamination cycle time, pathogen inactivation, and the maximum number of safe reuses.

In summary, we have demonstrated that the N95 facepiece shape and position within a UV-C decontamination system have substantial influence on the on-N95 UV-C dose distribution and concomitant decontamination efficacy. We introduce a workflow to combine optical modeling and *in-situ* quantitative PCI dosimetry to characterize on-N95 UV-C dose with high spatial resolution, high throughput, and near-ideal angular response. For the first time, we combined simultaneous and robust quantitative UV-C dose measurements with SARS-CoV-2 inactivation measurements at specific locations on N95 respirators to probe the relationship between on-N95 dose and pathogen inactivation within each UV-C exposure. The substantial variation in on-N95 UV-C dose and SARS-CoV-2 inactivation we observed in a single decontamination chamber highlights how nonuniform UV-C dose distribution impacts pathogen inactivation and total UV-C exposure (which influences N95 material degradation and the safe number of decontamination cycles). We further demonstrated that a lower-cost colorimeter accurately quantifies dose from PCIs, making the PCI quantification workflow more accessible. Additional investigation into alternative color metrics may extend the dynamic range of PCIs measured with lower-cost color readers. Future studies are needed to characterize SARS-CoV-2 dose response in more clinically relevant conditions, such as with the addition of soiling agents and on varying N95 models. Extending the dynamic range of PCIs, while maintaining a near-ideal angular response, is also critical for measurement of >∼200 mJ/cm^2^ UV-C dose on-N95. Overall, the on-N95 UV-C dosimetry approach here facilitates characterization of decontamination protocols of any UV-C system, supporting system-specific validation that is critical to ensuring safe and effective N95 decontamination.

## Data Availability

All relevant data are within the paper and the supplementary material. All reported data are available from the corresponding author upon reasonable request.

## Acknowledgments

We greatly appreciate the generous donation of the following materials: all UV-C chambers and bulbs (Betty Chang at Spectro-UV), handheld UV point lamp/source (Daniel Tristan at Spectroline), 3M 1860 N95s (Kristen Bole, Dr. Aenor Sawyer, and Dr. Nichole Starr at University of California, San Francisco) and Zemax OpticStudio Professional license (S. Subbiah and David Lineberger). We also thank the Streets Lab at University of California, Berkeley for kindly loaning the rotation platform (Thorlabs QRP02). We thank Douglas Fox for assistance with our Biological Use Authorization application, Brian Heimbuch for helpful discussion on viral inactivation measurements, as well as Csilla Timar-Fulep for assistance with Zemax Opticstudio.

We acknowledge the following funding sources: University of California, Berkeley College of Engineering Dean’s COVID-19 Emergency Research Fund (PI: A.E.H), Chan Zuckerberg Biohub Investigator Program (PI: A.E.H.), NIH training grant T32GM008155 (A.G. and A.S.), National Defense Science and Engineering Graduate (NDSEG) Fellowship (A.G.), National Science Foundation Graduate Research Fellowship Program (NSF GRFP, A.S.), University of California, Berkeley Siebel Scholarship (A.S.), and the Natural Sciences and Engineering Research Council of Canada (NSERC) postdoctoral fellowship (S.M.G). A.W.R. is an Open Philanthropy Fellow of the Life Sciences Research Foundation.

## Supporting Information

Figure S1. Two UV-C chambers have similar irradiance profiles over time and space

Figure S2. Equivalent performance of two ILT1254 radiometers

Figure S3. Template used to measure PCI color with Color Muse colorimeter

Figure S4. UV-C dose quantification from PCI color change

Figure S5. PCI calibration curve is batch-dependent

Figure S6. PCIs have near-ideal cosine angular response, measured from UV-C point source where power is independent of distance

Figure S7. Preprocessing of scanned N95 for optical model Figure S8. Map for *in-situ* measurements on chamber floor

Figure S9. Optical model identifies paired measurement sites for in-situ PCI measurements

Figure S10. Chamber floor map for on-N95 measurements

Figure S11. Simulation and *in-situ* measurements of UV-C distribution across chamber floor correspond.

Figure S12. *In-situ* irradiance mapping determines coupon placement

Figure S13. Chamber floor map for coupon study

Figure S14. SARS-CoV-2 inactivation does not depend on N95 expiration status

Figure S15. Correspondence between simulated and *in-situ* measured on-N95 UV-C dose distribution using PCIs

Figure S16. N95-to-N95 variation in morphology

Figure S17. Correspondence between radiometer and PCI-measured UV-C doses during coupon experiments

Figure S18. 0.5-1.5 J/cm^2^ UV-C yields >3-log inactivation of SARS-CoV-2 on N95 coupons

Figure S19. Chamber heating does not affect SARS-CoV-2 inactivation

Figure S20. 3M 1860 N95 coupons are hydrophobic

Figure S21. Normalized on-N95 UV-C dose-response curve for SARS-CoV-2 for 2 N95 facepieces

Table S1. Additional optical model specifications

Note S1. Generation of PCI calibration curves

Note S2. SARS-CoV-2 handling, inoculation, and TCID_50_ assay

Note S3. Assessing impact of chamber heating on SARS-CoV-2 viability

Note S4. In-process UV-C dose monitoring on chamber floor overestimates minimum on-N95 dose

