## Supplementary Material for "Nonuniform UV-C dose across N95 facepieces can cause 2.9-log variation in SARS-CoV-2 inactivation"

[#]These authors contributed equally.

**Figure S1.** Two UV-C chambers have similar irradiance profiles over time and space

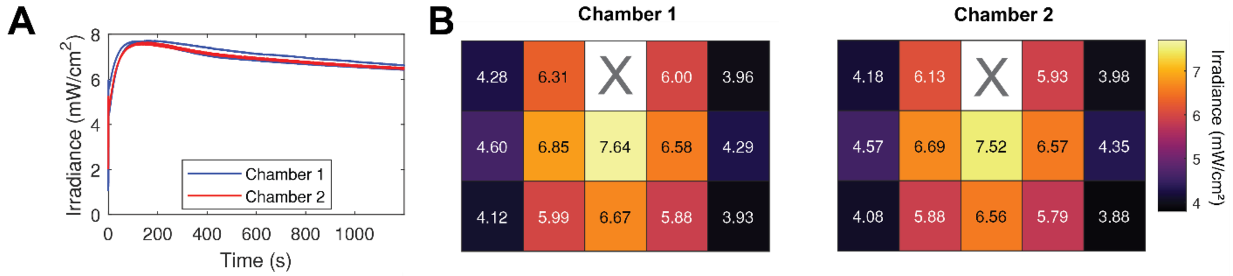

**Figure S1**. **Two UV-C chambers have similar irradiance profiles over time and space.** (A) Irradiance at the center of the chamber during the first 20 minutes of exposure after turning on the UV-C bulbs (N = 2 for each chamber). Note decrease in output over time after bulb warm-up. (B) Heatmaps of spatial irradiance distribution within each chamber (average of 3 replicate measurements at each location).

**Figure S2.** Equivalent performance of two ILT1254 radiometers

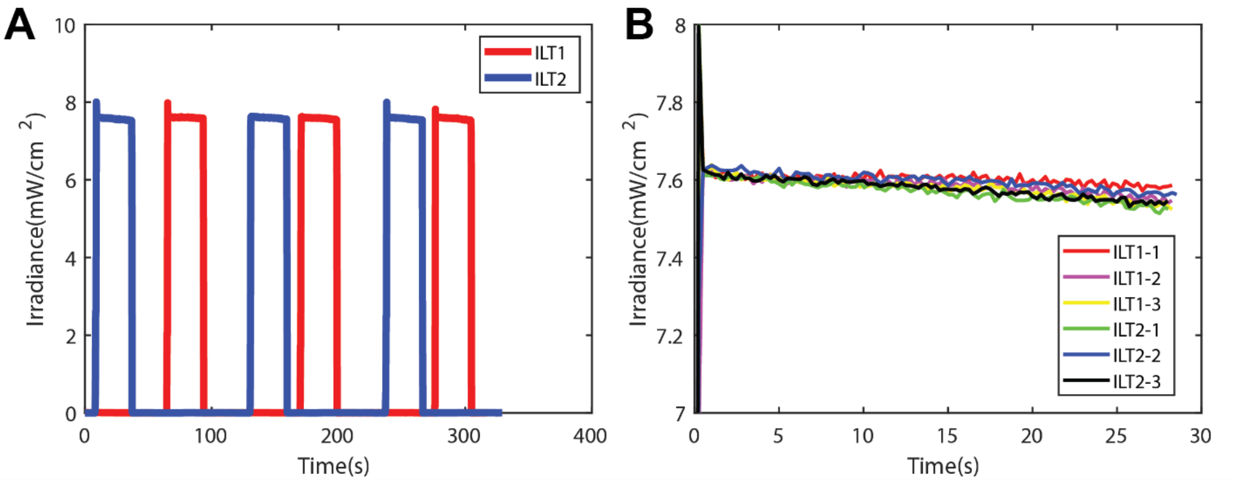

**Figure S2. Equivalent performance of two ILT1254 radiometers.** (A) Irradiance recorded by each radiometer (labeled ILT1, ILT2) when placed in the same location at the center of a UV-C chamber and exposed for 30 seconds in alternating fashion. (B) The irradiance data from (A) overlaid on top of one another, where t = 0 represents the start of each exposure.

**Figure S3.** Template used to measure PCI color with Color Muse colorimeter

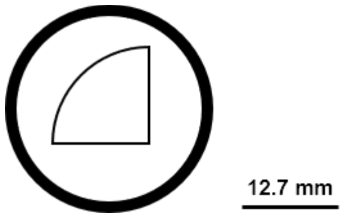

**Figure S3. Template used to measure PCI color with Color Muse colorimeter.** Because the Color Muse lacks a preview function, the template ensures that a quarter-circle PCI fills the Color Muse aperture.

**Figure S4.** UV-C dose quantification from PCI color change

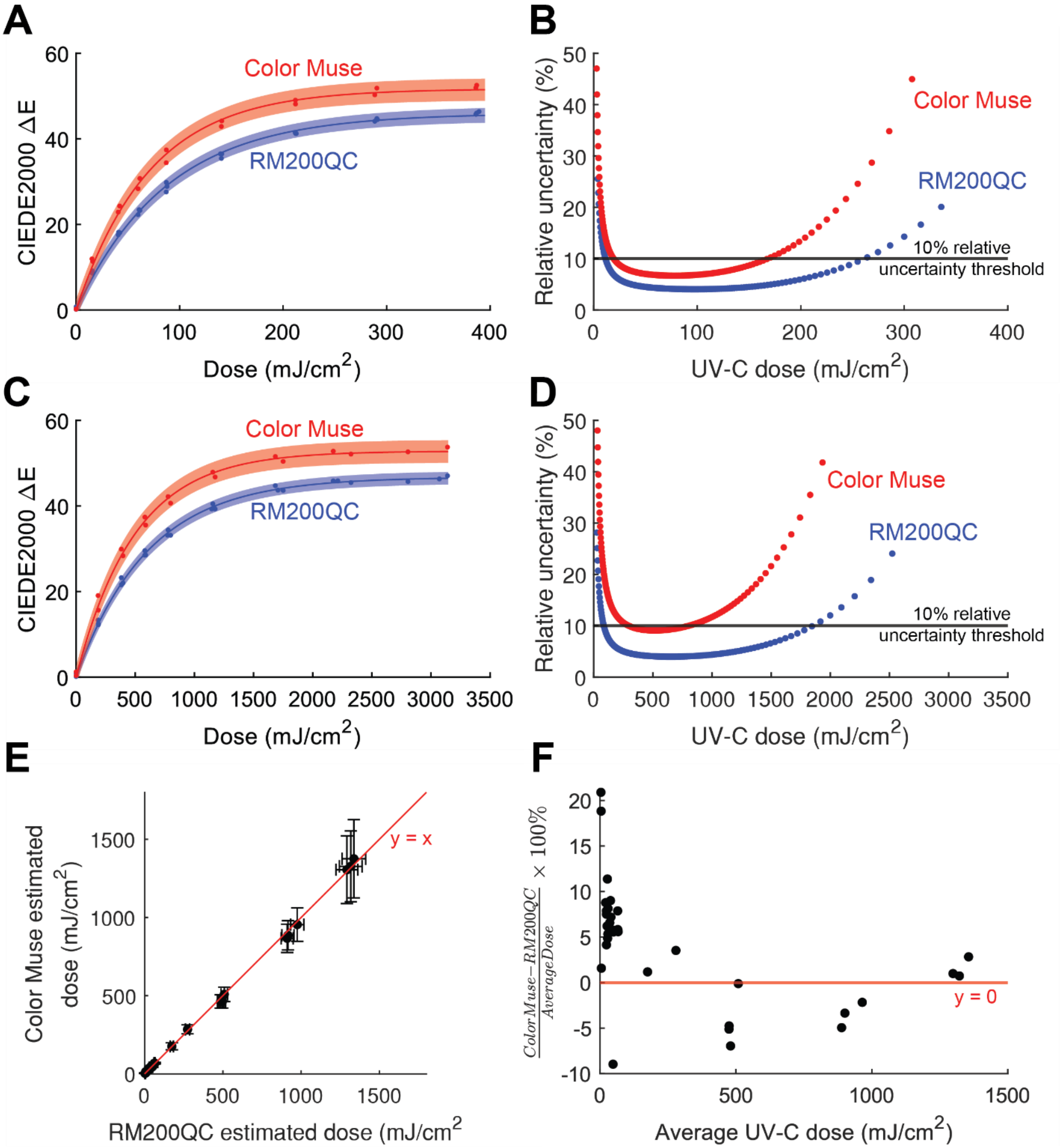

**Figure S4. UV-C dose quantification from PCI color change.** (A) Calibration curve relating UV-C dose to PCI color change (CIEDE2000 ΔE) measured with either the RM200QC spectrocolorimeter (N = 3 replicate data sets) or the Color Muse colorimeter (N = 2 replicate data sets). The shaded regions represent the 95% prediction interval on prediction of PCI color change from measured UV-C dose. Fit function is defined by Su & Grist, et al.[1] based on first-order reaction kinetics. For the RM200QC, $R^2$ = 0.9976, a = 46.0 (45.3, 46.7), b = 87.4 (83.6, 91.2). For the Color Muse, $R^2$ = 0.9963, a = 51.7 (52.8, 50.6), b = 71.2 (66.5, 75.9). (B) Relative uncertainty of dose measurement. Relative uncertainty is defined as half the width of the 95% confidence interval on UV-C dose measurements, divided by measured dose.

UV-C dose measurements have <10% relative uncertainty from 11.3 – 261.4 mJ/cm$^2$ (RM200QC) or 19.2 – 168.1 mJ/cm$^2$ (Color Muse). (C) Calibration curve of PCI covered by a 1.1 mm thick Borofloat glass attenuator, which extends the dynamic range. PCI color change is measured with either the RM200QC spectrocolorimeter (N = 3 replicate data sets) or the Color Muse colorimeter (N = 2 replicate data sets). For the RM200QC, $R^2$ = 0.9982, a = 46.7 (46.2, 47.3), b = 605.9 (584.3, 627.5). For the Color Muse, $R^2$ = 0.9960, a = 52.8 (51.7, 53.9), b = 495.6 (462.9, 528.3). To generate calibration curves of the PCI-Borofloat attenuator pair, the Borofloat was placed over the PCI during exposure but removed prior to PCI color measurement. The exposure times were also multiplied by a factor of $\frac{1}{\%T}$, where %T is the UV-C transmittance of the Borofloat glass, to account for the lower proportion of UV-C light reaching the PCI. For one batch of 1.1 mm-thick Borofloat glass, we measured a UV-C transmittance of 12.4% ± 0.4%, using a Spectroline XL-1500 chamber with BLE-1T155 bulbs and Model 308 data-logging UV radiometer with a 254 nm sensor (Optical Associates, Inc., OAI). (D) Relative uncertainty of dose measurements with 1.1 mm-thick Borofloat glass attenuator. UV-C dose measurements have <10% relative uncertainty from 85.0 – 1853.2 mJ/cm$^2$ (RM200QC) or 295.6 – 802.6 mJ/cm$^2$ (Color Muse). (E) Scatterplot of UV-C doses measured from the same PCI using either the RM200QC or ColorMuse. Error bars represent the uncertainty in the predicted dose, which arises from uncertainty in the calibration fit parameters and uncertainty in the ΔE measurement. (F) Scatterplot of the difference in dose measured by the ColorMuse and RM200QC. N = 34 PCIs.

**Figure S5.** PCI calibration curve is batch-dependent

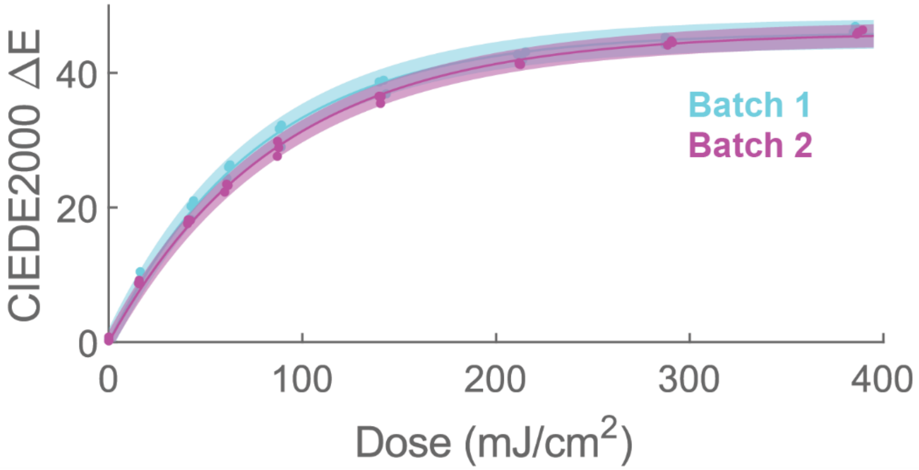

**Figure S5. PCI calibration curve is batch-dependent.** Each curve consists of 3 replicate datasets and was fit to the function based on first-order reaction kinetics defined by Su & Grist, et al.[1] For batch 1, $R^2$ = 0.9964, a = 46.0 (45.2, 46.8), b = 77.6 (73.5, 81.6). For batch 2, $R^2$ = 0.9976, a = 46.0 (45.3, 46.7), b = 87.4 (83.6, 91.2). Batch to batch variation may be due to changes in PCI starting color or variation in indicator reaction kinetics.

**Figure S6.** PCIs have near-ideal cosine angular response, measured from UV-C point source where power is independent of distance

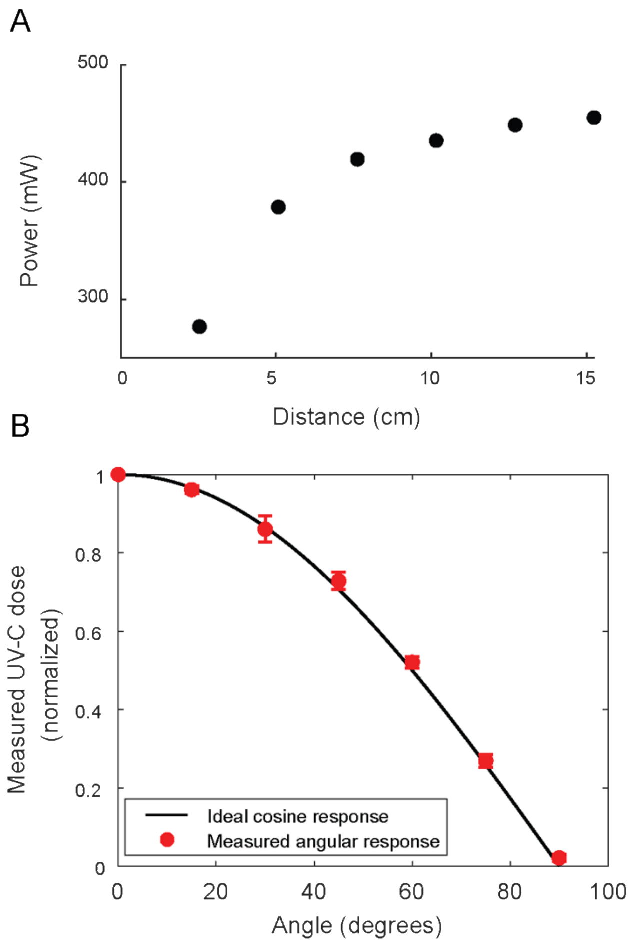

**Figure S6. Measuring the angular response of PCIs.** A) To determine where UV-C output power is independent of distance, irradiance was measured at several distances from the UV-C source using a radiometer. UV-C output power at each distance was calculated from the Keitz equation using average (N = 3) irradiance measured at different distances from the UV-C source[2]. The distance from the UV-C source at which output power changes by <5% between measurements (determined here to be ≥10.2 cm) was considered to be the point at which UV-C output power is independent of distance, which indicates that UV-C light is near-normally incident to a PCI perpendicular to the optical axis. B) PCIs have near-ideal cosine angular response. UV-C dose measured by a PCI with differing incident light angles from a UV-C point source, normalized to the measured dose at 0°. Angular response was measured between 0°-90° in 15° increments, in accordance with the range of angles used to characterize the angular response of other dosimeters.[3] Each PCI was affixed with double-sided tape to a glass microscope slide in a filter holder (Thorlabs FH2) mounted on a rotation platform (Thorlabs QRP02). PCIs were placed 10.2 cm away from a UV-C lamp (Spectroline E-Series handheld UV lamp with a BLE-2537S bulb and custom-built 2.54 cm diameter aperture). At this spacing, UV-C is near-normally incident to a PCI perpendicular to the optical axis based on UV-C output power measurements collected at different distances[2]. To ensure consistent UV-C output between exposures, a radiometer was used to monitor dose during each exposure;

all PCIs within an experiment were exposed to the same dose, as measured by the radiometer. To avoid shadowing, the radiometer was placed behind the PCI and at an offset such that the PCI/glass slide did not shadow the radiometer. The mean of 3 replicates is plotted (error bars are the standard deviation of the replicates).

**Figure S7.** Preprocessing of scanned N95 for optical model

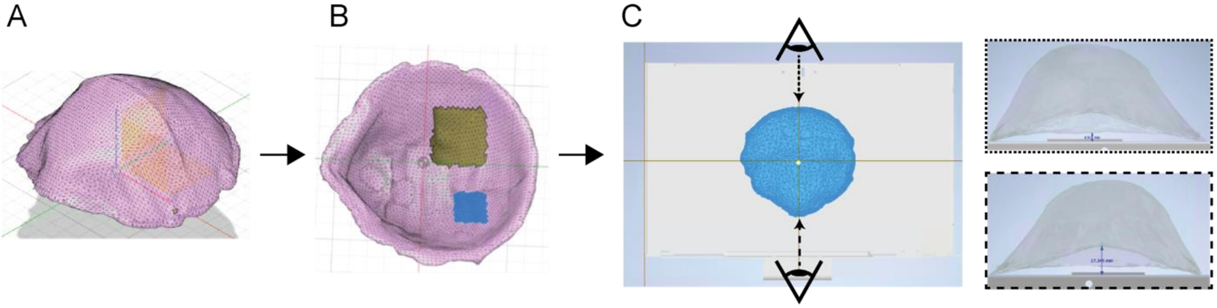

**Figure S7. Preprocessing of scanned N95 for optical model.** A) Scanned 3M 1860 N95 mesh model is roughly aligned to X-Y plane in Autodesk Fusion 360. B) The inner mesh layer facing the wearer was manually removed prior to converting the model to a solid and exporting as a .STEP file. C) A CAD model of the Spectronics XL-1000 (kindly provided by Spectro-UV) and modified N95 .STEP file were imported into an Autodesk Inventor assembly. The Spectrolinker-1000 was positioned to align the bottom of the back-left corner of the chamber with the origin. The model N95 was aligned close to the center of the top surface of the chamber floor. The pitch angle of the N95 was adjusted so that the height of the nosepiece and the chin piece approximated values measured *in situ*. The entire assembly was then imported into the optical modeling software for use.

**Figure S8.** Map for *in-situ* measurements on chamber floor

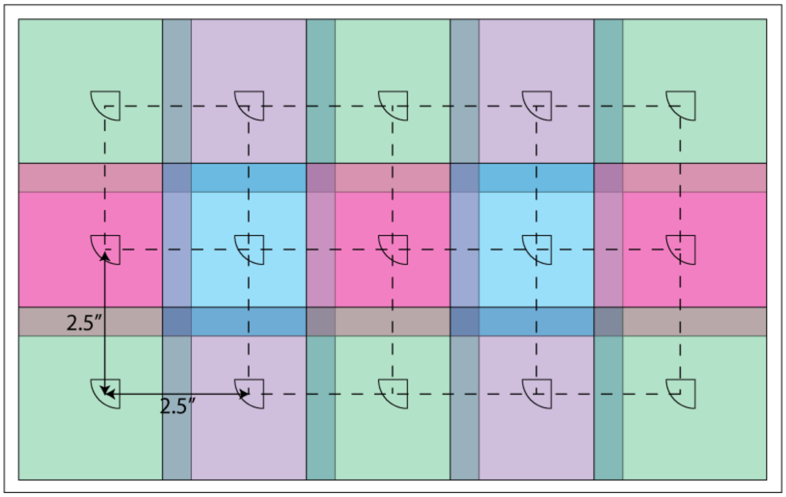

**Figure S8. Map for *in-situ* measurements on chamber floor.** Irradiance and dose were measured at 14 or 15 locations with a radiometer (colored squares) and PCIs (quarter-circles), respectively.

**Figure S9.** Optical model identifies paired measurement sites for *in-situ* PCI measurements

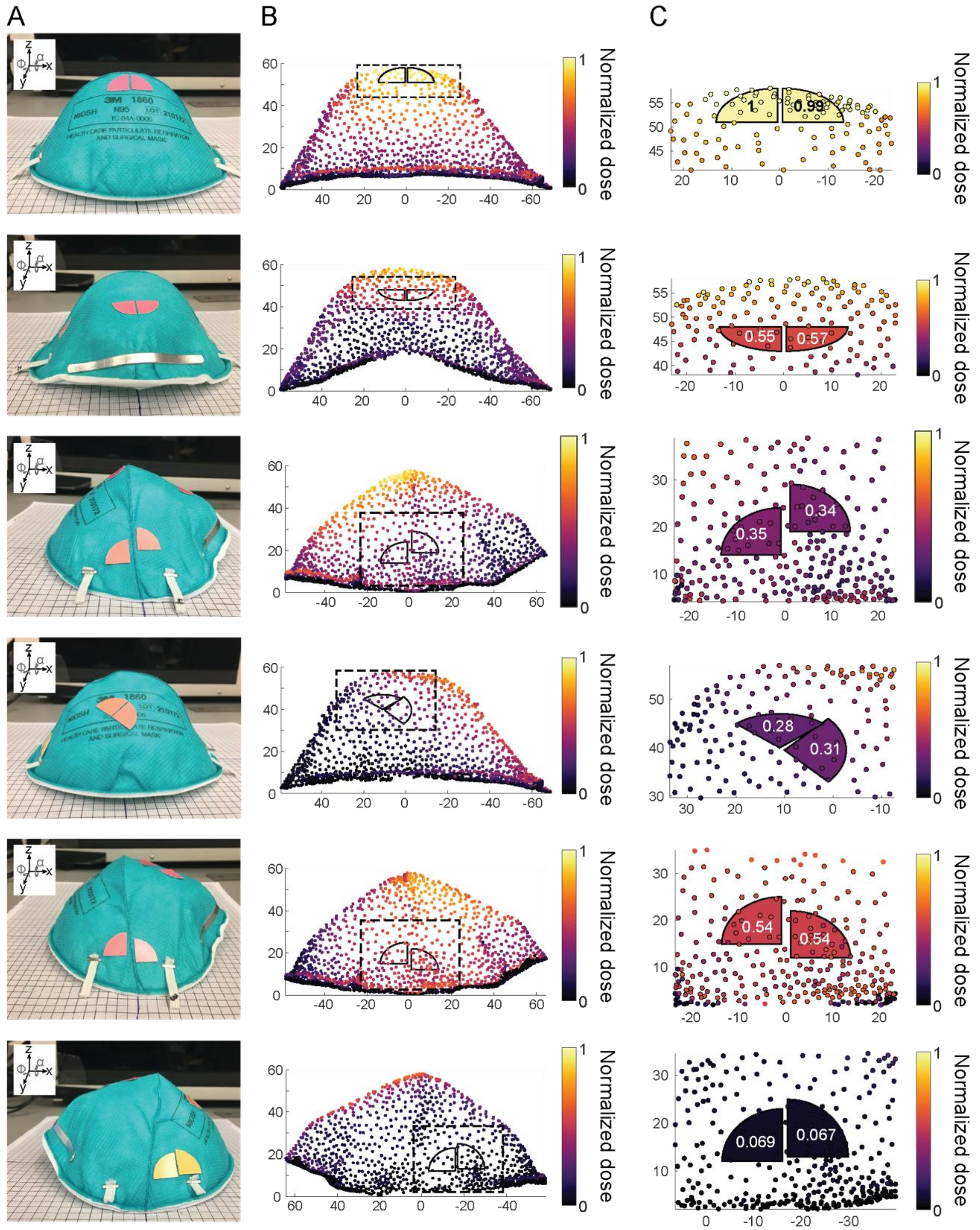

**Figure S9. Optical model identifies paired measurement sites for in-situ PCI measurements.** A) Images of PCIs placed on N95s taken post-UV-C exposure. While x-y-z axes are independent of view angle, rotational angles α and ϕ are defined relative to the view angle. B) Scatterplots of optical simulation output overlaid with outlines of PCIs estimated to receive similar doses, therefore identifying measurement sites. Dashed rectangular outline indicates area shown in C. C) High resolution plots of the PCIs and on-N95 simulation results from B projected onto a 2D plane. PCIs colored by average dose normalized to the maximum average PCI-measured dose, which is also indicated in overlying text. Axes in B) and C) show distance in mm. Due to N95 curvature, the PCI outline on the 2D projection may differ slightly from the true PCI footprint on an N95 *in situ*. We assume these differences are minimal.

**Figure S10.** Chamber floor map for on-N95 measurements

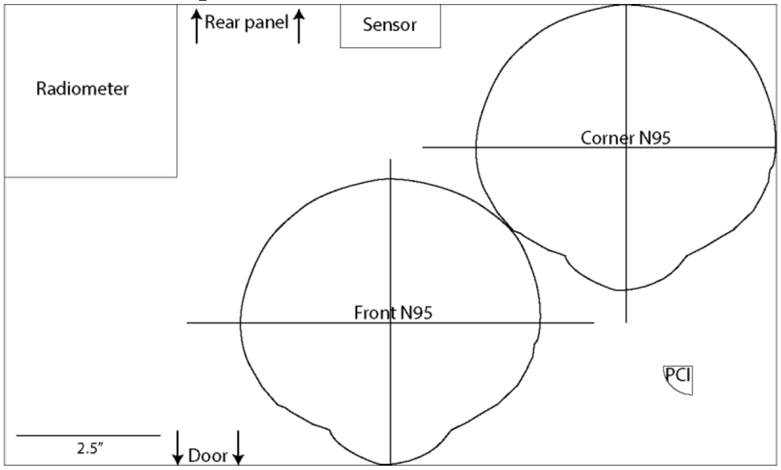

**Figure S10. Chamber floor map for on-N95 measurements.** Map positioned on chamber floor to ensure reproducible placement of all physical components for on-N95 measurements. "Sensor" indicates location of built-in sensor within the chamber.

**Figure S11.** Simulation correlates *in-situ* measurements of UV-C distribution across chamber floor

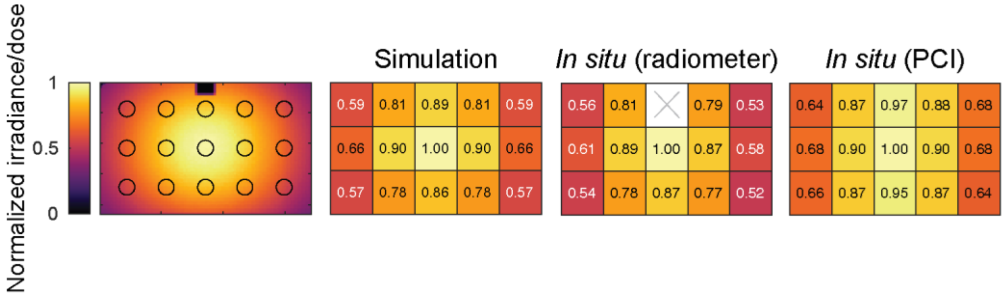

**Figure S11**. **Simulation correlates *in-situ* measurements of UV-C distribution across chamber floor correspond.** Spatial measurements across chamber floor estimated from simulation and measured *in situ* with a radiometer and PCIs. Leftmost heatmap shows locations from simulation from which values were averaged to compare to *in-situ* measurements. To map UV-C dose distribution on the chamber floor within the optical model, a rectangular detector with the surface area of the chamber floor was positioned at approximately the base height of the diffuser of the physical radiometer (35.175 mm). Small absorbing fiducials were introduced in the back left corner and above the built-in chamber sensor to assist with orientation during data post-processing. The average value within a 25.4 mm diameter circle (diameter of radiometer diffuser) at each *in-situ* position was determined using a custom MATLAB script.

Figure S12**.** *In-situ* irradiance mapping determines coupon placement

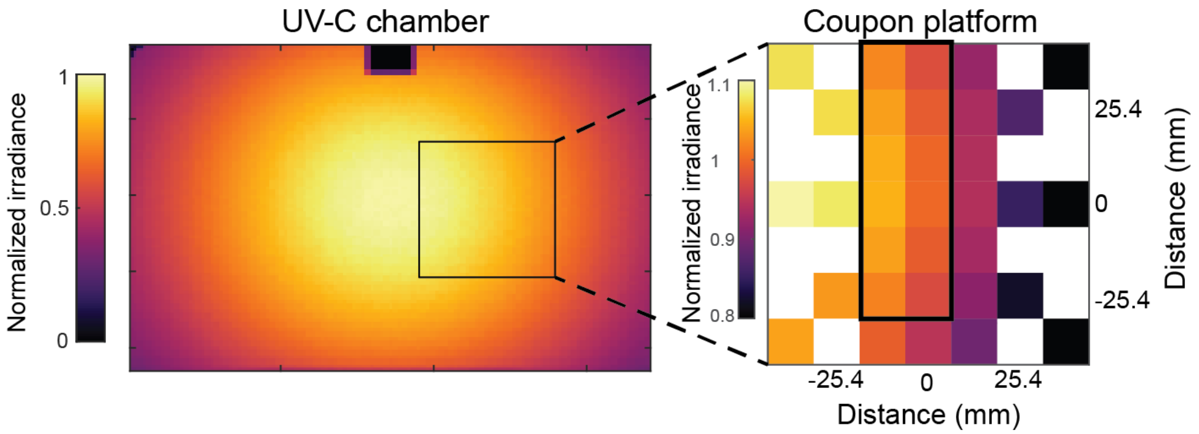

**Figure S12. *In-situ* irradiance mapping determines coupon placement.** *In-situ* irradiance mapping using the radiometer at the coupon platform location identifies a 25.4 mm × 63.5 mm region (black outline) where the irradiance varies < 10%. Within this area, 3 N95 coupons can fit for simultaneous exposure. Irradiance on the UV-C chamber heatmap is normalized to the maximum irradiance in the chamber. Irradiance on the coupon platform heatmap is normalized to the irradiance at the center (0 mm, 0 mm) position.

Figure S13. Chamber floor map for coupon study

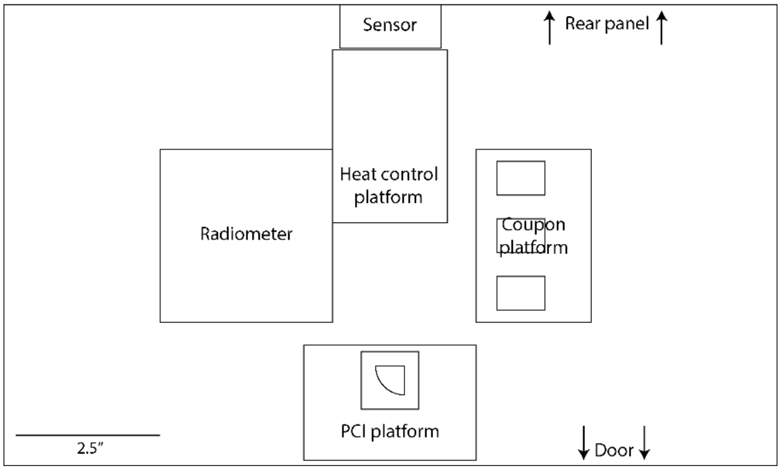

**Figure S13. Chamber floor map for coupon study.** Map positioned on chamber floor to ensure reproducible placement of all physical components involved in coupon study.

Figure S14. SARS-CoV-2 inactivation does not depend on N95 expiration status

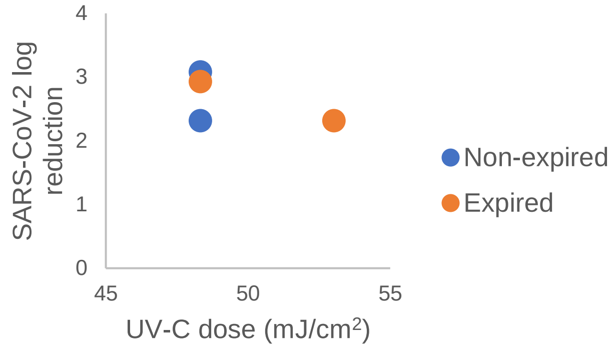

**Figure S14. SARS-CoV-2 inactivation does not depend on N95 expiration status.** No difference in SARS-CoV-2 UV-C response was observed between non-expired and expired 3M 1860 N95 material coupons. N = 2 replicates/condition.

Figure **S15.** Correspondence between simulated and *in-situ* measured on-N95 UV-C dose distribution using PCIs

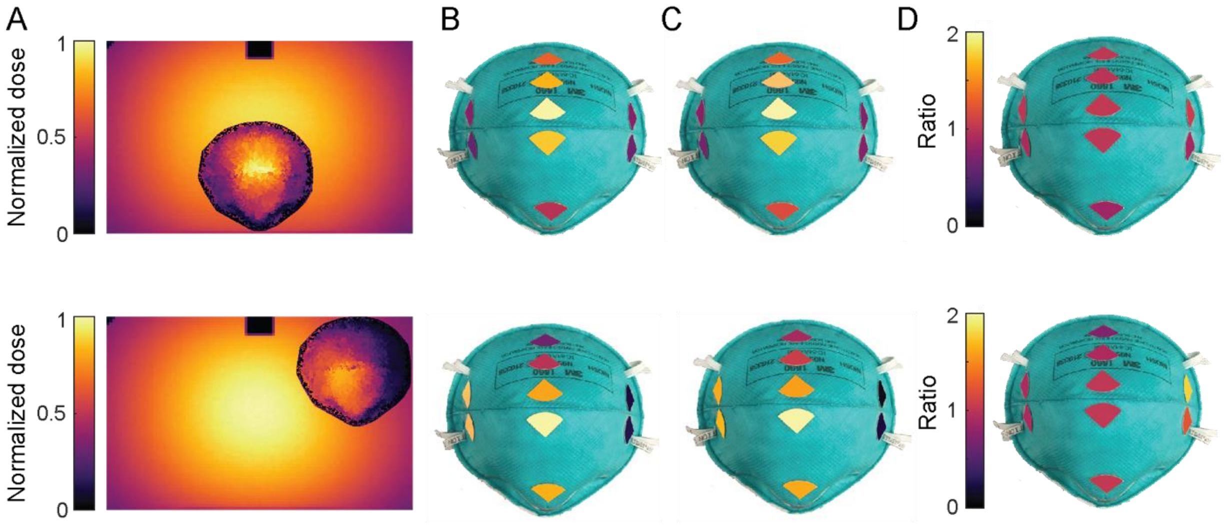

**Figure S15. Correspondence between simulated and in-situ measured on-N95 UV-C dose distribution using PCIs.** A) High resolution optical model output generated using Delaunay triangulation in Matlab (delaunayTriangulation) represented as heatmaps of UV-C dose across an N95 at the front-center of the chamber (top) and an N95 in the chamber corner (bottom). B) Normalized dose at PCI locations extracted from simulation results. C) In-situ normalized dose measured using PCIs (average of N = 3 replicates). D) Ratio of simulated to in-situ dose. All values were normalized to the highest on-N95 value, and on-N95 PCIs were false-colored to match color-mapped value. For PCI measurements, exposure times were chosen such that the on-N95 dose was within the dynamic range of the PCIs.

**Figure S16.** N95-to-N95 variation in morphology

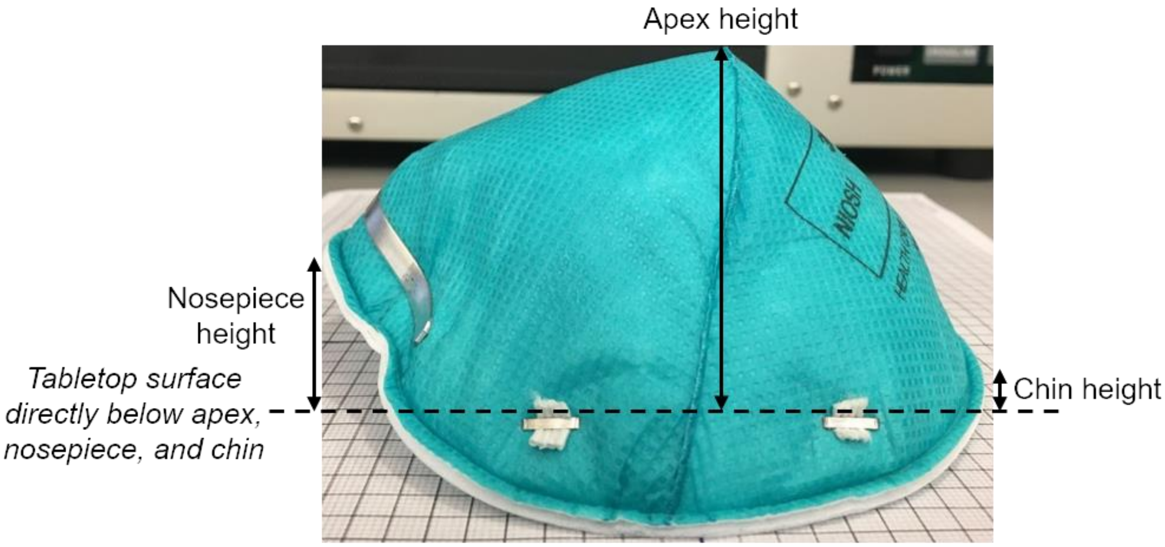

| N95 | Nosepiece height (mm) | Chin height (mm) | Apex height (mm) |
|---|---|---|---|
| Lot 1, N95 #1 *Used in simulation* | 16 | 5 | 58 |
| Lot 1, N95 #2 | 18 | 2 | 58 |
| Lot 2, N95 #1 | 19 | 7 | 61 |
| Lot 2, N95 #2 | 21 | 8 | 60 |
| Lot 2, N95 #3 | 22 | 5 | 61 |
| Lot 2, N95 #4 | 21 | 7 | 62 |

**Figure S16. N95-to-N95 variation in morphology.** Vertical heights from the tabletop to the seam on the nosepiece and chin seam of the N95, as well as to the apex. Heights are measured as the vertical distance from the nosepiece, apex, or chin to the tabletop directly below it.

**Figure S17**. Correspondence between radiometer and PCI-measured UV-C doses during coupon experiments

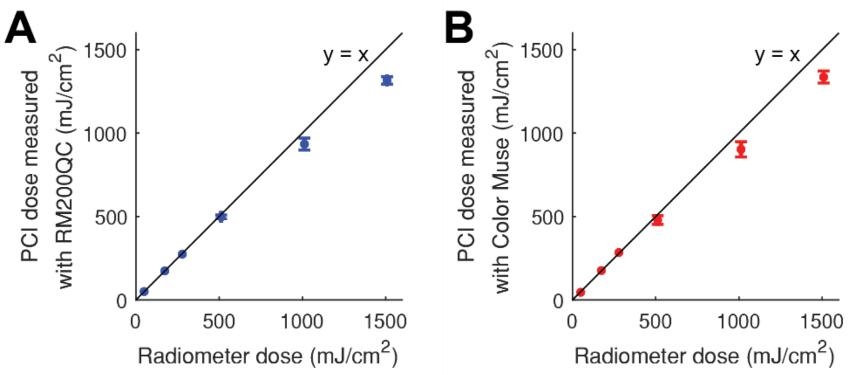

**Figure S17. Correspondence between radiometer and PCI-measured UV-C doses during coupon experiments.** PCI color change was measured with both the (A) RM200QC spectrocolorimeter, and (B) Color Muse colorimeter. For both color readers, N = 1 for radiometer doses <500 mJ/cm$^2$ and N = 3 for radiometer doses >500 mJ/cm$^2$. For dose measurements >168 mJ/cm$^2$, PCIs were coupled to a 1.1 mm-thick Borofloat attenuator. Vertical and horizontal error bars are the standard deviation of the estimated dose measurements. At UV-C doses <1000 mJ/cm$^2$, the PCI UV-C dose measurements were within 10% of the radiometer measurements. PCIs underestimated dose (compared to the radiometer) up to 13% at ~1500 mJ/cm$^2$, which may be due to the higher relative uncertainty in this dose range. Due to differences in temperature, humidity, and other environmental factors, the PCI response may also differ in the BSL-3 environment as compared to the non-BSL-3 location where PCI calibration curves were generated. Additionally, radiometer measurements were either made by integrating dose every 1 s in real-time or recording irradiance every 0.25 s and calculating integrated dose downstream; systematic differences between the two radiometer dose measurement methods may also contribute to the discrepancy between PCI and radiometer dose measurements.

Figure S18. 0.5-1.5 J/cm$^2$ UV-C yields >3-log inactivation of SARS-CoV-2 on N95 coupons

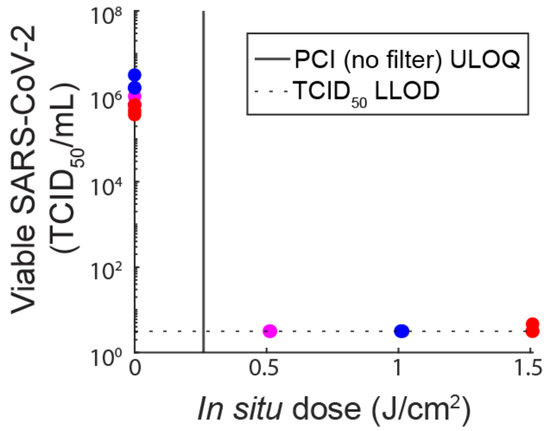

**Figure S18. 0.5-1.5 J/cm$^2$ UV-C yields >3-log inactivation of SARS-CoV-2 on N95 coupons.** ULOQ = upper limit of quantification. LLOD = lower limit of detection. Colors highlight temporally matched data (control coupons processed at the same time as exposed coupons). N = 3 replicates/condition

**Figure S19.** Chamber heating does not affect SARS-CoV-2 inactivation

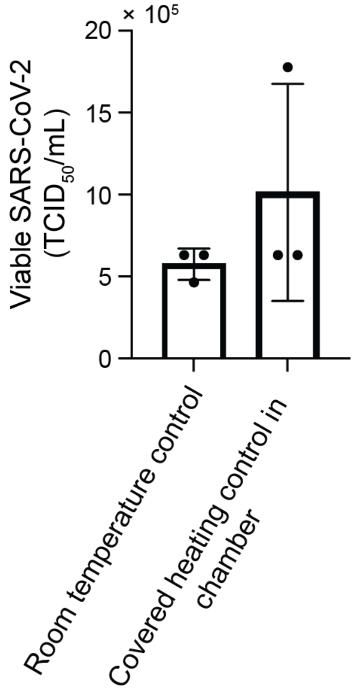

**Figure S19. Chamber heating does not affect SARS-CoV-2 inactivation.** Comparison of SARS-CoV-2 survival on N95 coupons at room temperature and within the chamber shielded from UV-C during illumination. The inoculated heating control coupon was placed on a platform of the same height as the coupon and PCI platforms, and in a location receiving approximately the same irradiance. An acrylic cover which was verified to block all UV-C was then placed on top of the heating control coupon, so that the coupon would be exposed to the temperature rise in the chamber but not to UV-C. A PCI was placed near the heating control coupon, under the acrylic cover, to verify that the heating control coupon is not irradiated with UV-C. $p > 0.9999$, Wilcoxon matched-pairs signed rank test.

**Figure S20.** 3M 1860 N95 coupons are hydrophobic

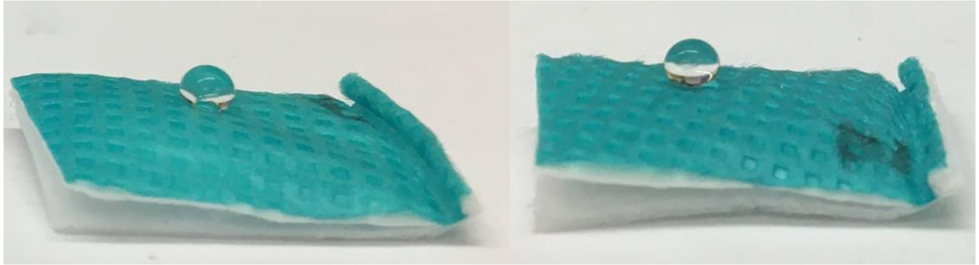

**Figure S20. 3M 1860 N95 coupons are hydrophobic**. A ~10 µL water droplet on expired (right) and non-expired (left) N95 coupons has a contact angle >90°, indicating high hydrophobicity. Additionally, the separation of the layers along the three sides without a seam may cause variable slope with respect to the UV-C source between coupons.

**Figure S21**. Normalized on-N95 SARS-CoV-2 UV-C dose-response curve for 2 N95 facepieces

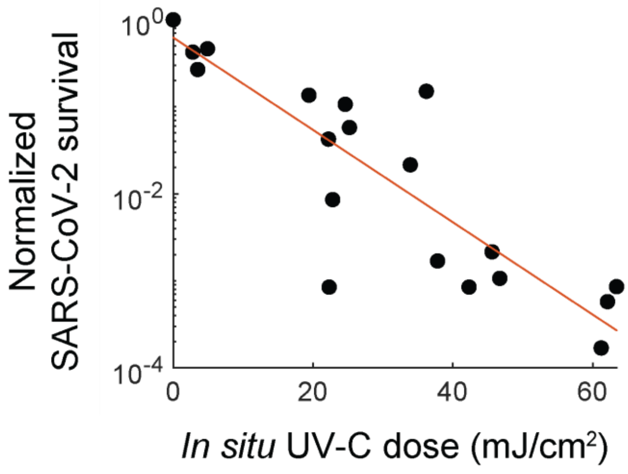

**Figure S21**. **Normalized on-N95 SARS-CoV-2dose-response curve for 2 N95 facepieces.** Normalized SARS-CoV-2 survival is calculated as $TCID_{50}$/mL divided by the time-matched negative control $TCID_{50}$/mL. Red line illustrates the linear regression on *in-situ* UV-C dose and log(survival) with equation: y = -0.0531*x - 0.2045 ($R^2$ = 0.78). Based on linear regression, the estimated $D_{90}$ dose is between 18.83 - 19.03 mJ/cm$^2$, depending on whether the y-intercept value is ignored or considered, respectively.

**Table S1.** Additional optical model specifications

| UV-C source information | | |
|---|---|---|
| Emission | 254 nm, monochromatic | |
| Number of UV-C bulbs | 5 | |
| Filament-to-filament length | 230 mm | |
| Diameter | 3 mm | |
| Rays/source tube during simulations | 5E7 | |
| **Reflective properties applied to chamber surfaces** | | |
| Component | % reflective | % diffuse scattering |
| Top reflector | 86 | 100 |
| Rear panel | 20 | 90 |
| Front door | 5 | 100 |
| Sides and bottom panel | 20 | 90 |

### **Note S1.** Generation of PCI calibration curves

PCI calibration curves, relative uncertainty, and dynamic range determination were performed as previously described[1]. Briefly, calibration curves relating PCI color change (CIEDE2000 ΔE) to received dose were established by placing a radiometer and a PCI at two locations of equal irradiance within the UV-C chamber.  PCIs were placed on a platform of similar height to the radiometer sensor (34 mm). The dose (calculated by integrating recorded radiometer irradiance over time) and CIEDE2000 ΔE measured after 9 different exposure lengths were fit to a function based on first-order reaction kinetics, as reported previously (a and b are fit parameters)[1]: $\Delta E \ = \ a(1 - e^{\frac{-dose}{b}})$ (Figure S4A,C). 95% confidence intervals (CIs) on UV-C dose measurements were calculated via propagation of error from standard deviations of the fit parameters and the standard deviation of replicate ΔE measurements of unexposed PCIs (N = 10 and 11 for RM200QC measurement of PCI batches 1 and 2, respectively; N = 6 and 5 for Color Muse measurement of PCI batches 1 and 2, respectively). Relative uncertainty was defined as $\frac{CI\ width}{2{*}estimated\ dose}$ (Figure S4B,D). The dynamic range was quantified as the dose range over which the relative uncertainty was <10%. Batch-specific calibration curves were generated due to batch-to-batch variability (Figure S5); calibration curves were also specific to the attenuator and colorimeter used. Custom MATLAB scripts performed PCI dosimetry analyses.

**Note S2.** SARS-CoV-2 handling, inoculation, and $TCID_{50}$ assay

*Virus preparation and stock titration:* SARS-CoV-2 stocks of the strain USA-WA1/2020 were obtained from the Biodefense and Emerging Infections (BEI) Research Resources Repository. Dulbecco's Modified Eagle Medium (DMEM, Sigma-Aldrich) containing 10% fetal bovine serum (FBS),100 U/mL penicillin, and 100 μg/mL streptomycin was used for all cell culture. To produce virus passage 1, SARS-CoV-2 stocks were amplified in Vero-E6 cells (ATCC® CRL-1586™). In brief, to generate passage 1, 50 μl of the BEI stock was inoculated onto confluent T-175 flasks of Vero-E6 cells and allowed to propagate until 50% cytopathic effect (CPE) was achieved (~48 hours post infection) at which time cells were lysed through 1 round of freeze and thaw. CPE was defined as any virus-induced cell death or change in cell morphology observed using brightfield microscopy. Supernatants were collected and clarified by spinning at 1500 rpm for 5 mins. The clarified viral supernatant was aliquoted and frozen at -80°C. Aliquots were thawed for production of virus passage 2, which was performed as above except using Calu-3 human lung epithelial cells (UC Berkeley Cell Culture Facility). The concentration of virus passage 2 stocks was assessed by 50% tissue culture infectious dose ($TCID_{50}$) assay using Vero-E6 cells and was determined to be 8 x $10^7$ $TCID_{50}$/mL. Passage 2 cells were used for all experiments.

*N95 facepiece/coupon inoculation:* All coupons or N95 facepiece viral measurement sites were inoculated by pipetting 3 aliquots of 16.67 μL, for a total of 50 μL, of passage 2 virus stock at $8 \times 10^7$ $TCID_{50}$/mL onto the N95 material. While most locations on the N95 facepiece can be inoculated, for the hydrophobic N95 model used in this study, we observed that beaded inoculation droplets would roll off of steeply sloped surfaces (e.g., base of the facepiece near the chin or nosepiece), precluding inoculation at some locations. Alternate droplet sizes or N95 orientations during drying may mitigate this challenge. The SARS-CoV-2 inoculation volume was selected to balance drying time and assay sensitivity. Inoculation sites were left to dry at room temperature for 3.5 hours in a biosafety cabinet. For paired on-N95 UV-C dose and SARS-CoV-2 inactivation measurements, where the PCIs were placed on the N95 prior to inoculation, we also verified that the N95s were not exposed to UV-C during the drying process. To do so, a PCI was positioned in the biosafety cabinet next to the N95 respirators while the inoculation sites dried, and the PCI color was measured using the RM200QC spectrocolorimeter before and after the 3.5 hour drying period to verify no change in color.

*Virus titration:* After irradiation, inoculated coupons or N95 facepiece measurement sites were extracted using 12mm biopsy punches. N95 facepiece punches were incubated in stationary 24-well plates containing cell culture media for ≥30 minutes. Viable SARS-CoV-2 virus was quantified by $TCID_{50}$ assay by incubating confluent Vero E6 cells in 96 well plates with 10-fold serial dilutions of viral extraction sample at 37°C/5% $CO_2$. Eight replicate wells were assessed per dilution. Five days after inoculation, CPE was scored visually under brightfield illumination using a 4×/0.13 NA objective. Wells with CPE exhibited either complete destruction of the cell monolayer, or large areas of cell lysis/cell debris. $TCID_{50}$ was calculated using the Reed-Muench method[4]. The limit of detection of the assay is 3.16 $TCID_{50}$/mL, which was determined by calculating the $TCID_{50}$ at which no CPE is observed in any replicate wells.

All study procedures were approved by the UC Berkeley Committee for Laboratory and Environmental Biosafety and conducted in agreement with BSL-3 requirements.

### **Note S3.** Assessing impact of chamber heating on SARS-CoV-2 viability

To investigate whether heating within the chamber during treatment contributes to SARS-CoV-2 inactivation, we first monitored the chamber temperature during UV-C exposures. Temperature in the UV-C chamber was recorded in preliminary experiments (outside of BSL-3) over time using a USB temperature/RH datalogger placed at the center of the chamber (Digi-Sense UX-20250-42). After UV-C bulb warm-up, we measured a chamber temperature of ~27 $^{o}$C. Over an exposure time of 200 s, we observe a temperature increase of 0.016 ± 0.001 $^{o}$C/second (N = 3 exposures), which corresponds to <3.3 $^{o}$C increase over the total cumulative exposure time of all replicates (183 seconds). Thus, we do not anticipate the total temperature increase to contribute to SARS-CoV-2 inactivation, as equivalent SARS-CoV-2 survival after 30 minutes at 22°C and 37°C has been observed[5].

Furthermore, to directly verify that heating in the UV-C chamber did not contribute to SARS-CoV-2 inactivation, we measured SARS-CoV-2 inactivation on inoculated N95 coupons shielded from UV-C inside the chamber during exposures. An additional inoculated 'heating control' coupon was included in the chamber under UV-C-blocking material during the 175, 300, and 500 mJ/cm$^2$ exposures of the dose-response characterization on N95 coupons. Like the exposed and room-temperature unexposed control coupons, the inoculation site on the heating control coupon was excised and processed immediately after each exposure. Compared to control coupons kept outside of the chamber during exposures, we observed no significant difference in viable SARS-CoV-2 TCID50/mL (N = 3 replicates, p>0.9999, Wilcoxon matched-pairs signed rank test, Figure S19). These observations suggest that chamber heating does not contribute to SARS-CoV-2 inactivation, as supported by literature on SARS-CoV-2 stability at measured chamber temperatures.

**Note S4.** In-process UV-C dose monitoring on chamber floor overestimates minimum on-N95 dose

To assess whether dose measured at a location off-N95 could be used for in-process dose monitoring of decontamination cycles, we characterized the relationship between on-N95 dose and the dose measured on at a specific location on the chamber floor. In-situ dose is often monitored at an off-N95 location in decontamination protocols[6], as on-N95 dose measurements would shadow the underlying N95 region from irradiation. To test whether UV-C dose monitoring on the chamber floor could serve as a proxy for the lowest dose received by N95s in the chamber, we compared the dose received in two of the lowest-dose locations on the chamber floor to the lowest dose measured on-N95.

Based on the simulation of UV-C dose distribution across the chamber floor (Figure 3A,C), we anticipated that the corners of the UV-C chamber receive the lowest on-floor dose, and thus we chose to measure dose at two corners using a radiometer and PCI. Compared to the maximum on-N95 dose measured, the doses on the floor in the chamber corners were 49.5% ± 1.6% (radiometer location) and 44.0 ± 0.7% (floor PCI location) of the maximum on-N95 dose, whereas the lowest on-N95 dose measured was 6.0% ± 1.6% of the maximum on-N95 dose (Figure 3C,D). Thus, in the UV-C chamber tested here, dose monitoring on the chamber floor cannot serve as a proxy for the lowest on-N95 UV-C dose, even if on-floor dose is monitored in the lowest-irradiance locations. As can be seen in the heatmaps and values reported in Figure 3C and 3D, steeply sloped regions (particularly on the corner N95) receive several-fold lower dose than the lowest-irradiance location on the chamber floor. If a protocol is tuned only to ensure the on-floor monitoring location receives sufficient dose for decontamination, the N95s will not be fully decontaminated. Instead, care should be taken to quantify the relationship between the lowest on-N95 UV-C dose and the dose received at any in-situ monitoring point. This relationship can then be used to ensure that all N95 surfaces receive at least the on-N95 target dose, as described previously[1].
